# Multi-omics models for hypertension prediction using blood pressure PRS and omics scores

**DOI:** 10.64898/2026.11.13.26368055

**Authors:** Ziqing Wang, Yana Hrytsenko, Sivan Almogy, Brian W Spitzer, Yogesh Purushotham, Kent Taylor, Henry J Lin, Usman Tahir, Laura M Raffield, Stephen S Rich, Jerome I Rotter, Zsu Zsu Chen, Tamar Sofer

## Abstract

**Background:** Hypertension is a major modifiable risk factor for cardiovascular disease, yet the relative contributions of proteomic (ProtS), metabolomic (MetS), and polygenic risk scores (PRS) to incident hypertension risk remain incompletely characterized.

**Methods:** We developed ProtS and MetS for systolic (SBP) and diastolic blood pressure (DBP), and hypertension using Elastic Net regression in the Multi-Ethnic Study of Atherosclerosis (MESA). We incorporated these scores with previously developed BP PRSs into 5 incremental XGBoost models, with external validation in the Jackson Heart Study (JHS). We further applied transfer learning to adapt the MESA-derived models to JHS. Elastic Net selected proteins were prioritized using Mendelian randomization, colocalization, and functional analyses.

**Results:** In MESA, model discrimination improved with the addition of omics scores, with the combined model achieving an area under the curve (AUC) of 0.72 compared with 0.63 for the clinical baseline model. In JHS, ProtS showed the highest AUC of 0.68 and the smallest cross-cohort performance decline, compared to MetS and PRS. Incorporating measured SBP and DBP substantially improved discrimination, with the PRS+BP model achieving an AUC of 0.80 in MESA and the baseline+BP model achieving an AUC of 0.74 in JHS. Among JHS participants with normal baseline BP, ProtS provided the strongest discrimination among models without measured BP (AUC = 0.62), while PRS+BP model achieved an AUC of 0.69. Leaf-weight refreshing consistently improved calibration in JHS while preserving discrimination. Mendelian randomization identified 77 proteins associated with BP traits or hypertension, involved in vascular, inflammatory, renal, and natriuretic pathways.

**Conclusions:** ProtS demonstrated superior predictive performance and cross-cohort transferability among the omics modalities evaluated, while measured BP remained the strongest predictors of incident hypertension. Proteomic and genetic information may provide complementary biological signals for identifying individuals at elevated hypertension risk, particularly before clinically elevated BP is detectable.

## Introduction

Hypertension affects over 1 billion adults worldwide and is the leading modifiable risk of cardiovascular disease mortality, imposing a substantial public health burden due to its high prevalence^1,2^. This burden is especially pronounced in non-Hispanic Black populations, which have the highest age-adjusted prevalence of elevated blood pressure (BP) and stroke mortality in the US and globally^3^. Early risk stratification of high-risk individuals is central to preventing hypertension and the downstream target organ damage, as BP elevation is largely asymptomatic^4^.

The hypertension etiology is influenced by complex interactions between genetic predisposition and life-course exposures, with estimated heritability ranging from less than 10% to over 60%^5,6^. Polygenic risk scores (PRS) aggregating variants identified by hypertension and BP GWAS show consistent but modest associations with incident hypertension^7–9^, with performance varying by race/ethnic background due to allele frequency heterogeneity and linkage disequilibrium (LD) differences^10^. Beyond PRS, large-scale omics technologies have opened new opportunities for understanding hypertension biology. For example, omics risk scores have been developed to represent health outcome status and are useful for studying disease etiology^11^. BP associated metabolic signatures including levels of cortisol, tryptophan metabolites, lipid species, as well as dietary metabolic patterns, have explained meaningful BP variance across cohorts^12–14^, suggesting their utility as biomarkers integrating environment-related HTN risk. Large-scale proteomic studies have similarly identified BP-associated proteins involved in inflammatory, vascular remodeling, renal, and extracellular matrix pathways^15–17^. Multi-omics integration, especially metabolomics and proteomics, has been shown to improve cardiovascular prediction beyond individual platforms^18,19^, motivating the combination of complementary biological dimensions to improve disease classification.

These developments underscore the need for analytical approaches capable of integrating diverse, high-dimensional data and modeling complex relationships among risk factors. Machine learning (ML) methods have improved cardiovascular risk prediction by capturing non-linear relationships and feature interactions, with interpretable feature importance through SHapley Additive exPlanations (SHAP) values^20^. However, prediction models trained in one population often show degraded performance when applied to independent populations due to limited generalizability^21^. In this study, we developed ML-based prediction models for incident hypertension incorporating PRS, metabolomic (MetS) and proteomic scores (ProtS), using a multi-ethnic discovery cohort and evaluated their performance in an independent external validation cohort predominantly comprising Black-American participants. To address cross-cohort generalization, we implemented transfer learning strategies that adapted the models using a fraction of data from the target population. In addition, proteome-wide Mendelian Randomization (MR) was performed to identify proteins showing potential causal effects on hypertension. This integrated framework aims to systematically evaluate the relative contributions of genetic, proteomic, and metabolomic modalities beyond established clinical risk factors for incident hypertension, and advance mechanistic understanding of hypertension biology.

## Methods

The primary study design is illustrated in Figure 1. ProtS and MetS for BP traits were first developed using Elastic Net regression in Multi-Ethnic Study of Atherosclerosis (MESA) participants, and constructed in The Jackson Heart Study (JHS) as weighted sums of selected proteins (ProtS) or metabolites (MetS). Together with previously developed PRSs for BP traits, the predictive utility of the three omic score types for incident hypertension was examined using a train-validation XGBoost framework across the two prospective cohorts. Specifically, to develop models that are generalizable across cohort populations, prediction models were trained in MESA and subsequently evaluated in the independent JHS cohort (Figure 1). Participants with prevalent hypertension at baseline were excluded, yielding, in MESA, analytic samples of 2,562 participants for each of the three models: the baseline model, PRS model, and all-scores model, 2,511 for MetS model, and 2,301 for ProtS. The corresponding analytic sample sizes in JHS were 1,187, 852, and 398 participants, respectively (Figure 1). We also performed a separate MESA-specific analysis splitting the data internal training (80%) and validation (20%) datasets to assess the potential limitations of model generalizability due to cohort heterogeneity (Figure 1).

**Figure 1.**
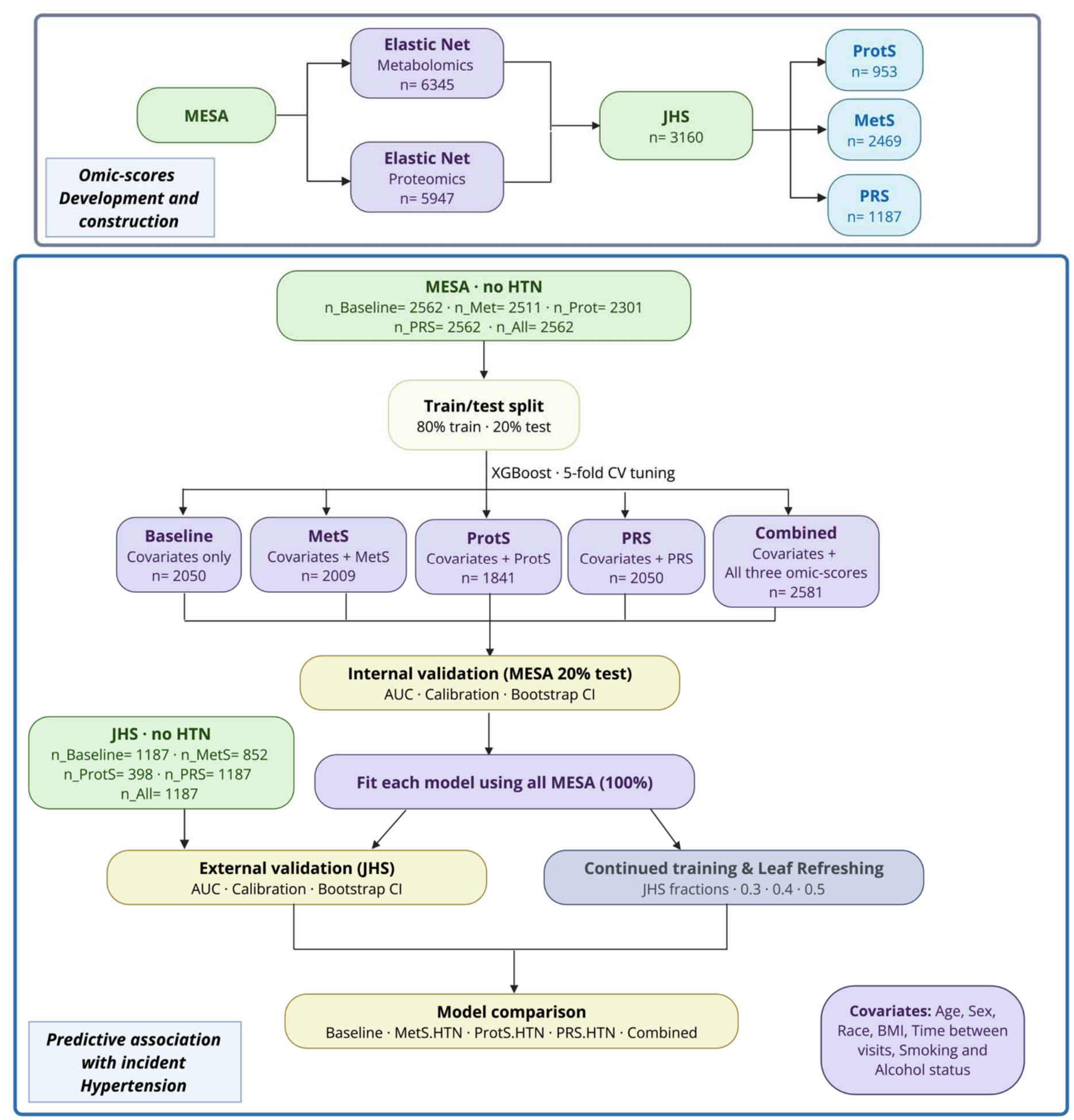
Study design overview: omic-score development and predictive modeling pipeline for incident hypertension. Top panel: Elastic net regression models were trained on MESA metabolomics and proteomics data to generate weights for the metabolomic (MetS) and proteomic scores (ProtS), respectively, which were then applied in JHS to derive ProtS and MetS. Polygenic risk score (PRS) was constructed based on previously published GWAS results. Bottom panel: 80/20 train/test split was applied among MESA participants free of hypertension at baseline, to develop five XGBoost models: Baseline, MetS, ProtS, PRS, and Combined (all three omic-scores), with covariates including age, sex, race, body mass index (BMI), time between visits, smoking, and alcohol status. Models were first assessed via internal validation on the MESA 20% test set, then refit using 100% of MESA before being applied to JHS participants free of hypertension. In addition to external validation of the MESA-fit models, continued training and leaf refreshing were performed using three training fractions of JHS data.

### The Multi-Ethnic Study of Atherosclerosis (MESA) Study

MESA is a longitudinal, communitybased cohort that enrolled 6,814 participants aged 45–84 years at baseline (Exam 1; 2000-2002) from six U.S. field centers (Baltimore, MD; Chicago, IL; Forsyth County, NC; Los Angeles, CA; New York, NY; and St Paul, MN)^22^. All participants were free of overt clinical cardiovascular diseases at enrollment. Standardized questionnaires were used to collect self-reported sociodemographic information, including age, sex, race/ethnicity (White, African American, Hispanic, and Chinese American)^22^. Hypertension was defined as systolic blood pressure (SBP) ≥140 mmHg, diastolic blood pressure (DBP) ≥90 mmHg, current use of antihypertensive medication, or a self-reported physician diagnosis^23^. To estimate BP among participants receiving antihypertensive therapy, 15 mmHg and 10 mmHg were added to the measured SBP and DBP values, respectively, following a previously described approach ^24^.

### The Jackson Heart Study (JHS)

JHS is a community-based, observational cohort comprising 5,306 non-institutionalized African American adults ≥21 years of age from the Jackson, Mississippi metropolitan area between 2000 and 2004^25^. At each clinic visit, participant completed standardized questionnaires to provide information on sociodemographic, lifestyle, and family medical history^26^. Hypertension was defined using the same criteria as in MESA at each visit, and SBP and DBP measurements were similarly adjusted for medication use.

### Proteomic profiling

Proteomic profiling was performed using the Olink Explore 3K and 5K platform (Olink Proteomics, Uppsala, Sweden), which quantified 2941 and 5420 protein assays^27,28^, respectively. Briefly, paired oligonucleotide-conjugated antibody probes selectively bind to target proteins. Upon target binding, the antibody-conjugated oligonucleotides hybridize and undergo a proximity-dependent DNA polymerization reaction, generating a unique DNA template that is subsequently quantified using a microfluidic real-time PCR platform (Biomark HD, Fluidigm). Data quality was ensured through Olink’s standard quality control procedures, including within-run and interplate normalization. Protein measurements were further normalized across plates to reduce residual batch effects and subsequently rankbased inverse normal transformed to have an approximate mean of 0 and standard deviation of 1. In total, 5947 MESA and 287 JHS participants had protein data profiled using Olink 3K platform, with additional 1381 JHS participants profiled using Olink5K platform from their respective baseline exams.

### Metabolomic profiling

For Both MESA and JHS, metabolomic profiling was performed on fasting plasma samples collected at baseline using liquid chromatography–tandem mass spectrometry (LC–MS/MS) platforms. Polar metabolites and lipids were captured through hydrophilic interaction liquid chromatography (HILIC) operated in positive and negative ionization modes, and reversed-phase C8 chromatography, respectively ^23,29,30^. Water-soluble and lipid metabolites were quantified on a Nexera X2 Ultra-High Performance Liquid Chromatography (U-HPLC) system (Shimadzu Corp., Marlborough, MA) coupled to a Q Exactive mass spectrometer (Thermo Fisher Scientific, Waltham, MA)^29^. Cross-batch normalization was applied to all samples, and coefficients of variation metrics were computed to assess data quality^22^11^29^. Both targeted and untargeted spectral features were subsequently processed using HILIC chromatography coupled to a Thermo ID-X mass spectrometer (Thermo Fisher Scientific, Waltham, MA) to construct a comprehensive MS/MS spectral library for metabolite identification, as previously reported^22^.

In this study, 663 metabolites annotated with Human Metabolome Database (HMDB) identifiers were used for metabolite score development in 6345 MESA participants from baseline. In contrast, only 109 metabolites were annotated with HMDB ID in 2469 JHS participants due to lack of C8/C18 lipid data collection. For metabolites measured on multiple LC-MS methods, duplicate measurements were resolved by retaining the measurement from the LC-MS method with the lowest coefficient of variation, calculated within each method. Metabolites with more than 25% missing values across participants were excluded. In metabolites with less missingness, missing values were imputed using the half-minimum method^31^, followed by rank-normalization to a standard normal distribution for each metabolite.

### Whole genome sequencing data and PRS construction

Whole-genome sequencing (WGS) data were obtained from the Trans-Omics for Precision Medicine (TOPMed) Program Freeze 10b release. DNA samples were sequenced through the TOPMed program and the National Human Genome Research Institute (NHGRI) Genome Sequencing Centers, and variants were jointly called using the TOPMed Informatics Research Center pipeline as described in the TOPMed Freeze 10 WGS Methods documentation (https://topmed.nhlbi.nih.gov/topmed-whole-genome-sequencing-methods-freeze-10). Sequencing reads were aligned to the GRCh38 reference genome using the functionally equivalent alignment pipeline. Genetic Principal Components (PCs) and kinship coefficients were computed centrally using the PC-Relate and PRAiR algorithms implemented in the GENESIS R package (v2.30.0)^32^. WGS data were available for 6290 MESA and 3160 JHS participants.

For PRS construction, we retained only variants that passed TOPMed quality control (FILTER = PASS, quality score = 256), and had a minor allele frequency ≥ 0.01 in the TOPMed dataset used in this study. SBP, DBP and hypertension PRSs were calculated by PLINK^33^ based on previously published PRS weights^34,35^ obtained from the Polygenic Score (PGS) catalog^36^ (PGS003968 for SBP, PGS003964 for DBP, and PGS004234 for hypertension). In particular, the SBP and DBP PRSs were developed as weighted sums using multiple independent genome-wide association study (GWAS) summary statistics^35^ (Table S1). The hypertension PRS was derived as the unweighted sum of PRSs for SBP and DBP from the Million Veteran Program (MVP), and a hypertension-specific PRS from a pan-ancestry GWAS conducted in the UK Biobank^34^ (Table S1).^33^

### Statistical analyses

#### Protein and metabolite score development for blood pressure traits in MESA

We developed ProtS and MetS to capture baseline molecular signatures of BP. We used all available MESA participants for score derivation and reserved JHS as the external cohort for evaluating performance. Specifically, all available MESA participants with proteomic (n= 5947) and metabolomic data (n= 6345) were used to develop ProtS and MetS for SBP, DBP, and hypertension via Elastic Net regression implemented in R package glmnet (v4.1.8) with 10-fold cross-validation to select the best regularization parameters. BP measures were from the same time point as the drawn blood used for measuring metabolites and proteins. MetS were derived by regressing rank-normalized metabolites against SBP, DBP, and hypertension status separately, while adjusting for age, sex, BMI, and race/ethnicity. ProtS were derived analogously with an additional filtering step. Specifically, proteins were first selected for inclusion based on their associations (p-value < 0.05) with baseline hypertension and SBP/DBP measurements, using logistic regression for hypertension and linear regression for SBP and DBP, respectively, while adjusting for the same covariates above. This independent filtering step was applied because the large number of features relative to sample size could reduce performances of penalized regression methods^37^. ProtS and MetS were subsequently calculated as the weighted sum of the selected features with non-zero Elastic Net coefficients, representing the linear combination of features jointly most predictive of each trait. The BP-related omic scores and their corresponding baseline phenotypes were validated using the linear models for SBP and DBP, and logistic regression for hypertension, adjusting for age, sex, and BMI in the full JHS dataset. In sensitivity analysis, we also developed and evaluate PortS and MetS in MESA, using a 80-20% training-evaluation dataset split.

#### Development of gradient boosting trees models for incident hypertension in MESA

Incident hypertension was modeled as binary outcome using gradient boosted decision trees implemented in the XGBoost framework using R package xgboost^38^ (v1.7.11.1), with 5-fold cross-validation for hyperparameter tuning performed using the caret^39^ package (v.7.0.1). Five prediction models with progressively increasing levels of complexity were developed to evaluate the incremental contributions of omics and genetic information to hypertension risk prediction: 1) a baseline clinical model including age, sex, self-reported race, BMI, smoking status, alcohol use, and time between study visits; 2) a MetS model adding MetS_SBP, MetS_DBP, and MetS_HTN (hypertension) to above clinical variables; 3) a ProtS model including the three BP ProtS and clinical features; 4) a PRS model including the three BP PRSs, 5 genetic PCs and clinical features; and 5) an all-scores model including all clinical, omics and genetic scores in a single model (Figure 1). In primary analysis, we used the ProtS and MetS developed in the full MESA dataset despite potential overfitting, in order to use exactly the same variables in MESA and subsequent JHS analysis. An African American background (Race-AFR) variable was created and assigned a value of 1 for all JHS participants to match the self-reported race variable used in the MESA-trained model. In sensitivity analysis (described later), we also used ProtS and MetS developed only on the MESA training dataset. Although XGBoost is robust to correlated predictors, measured SBP and DBP were excluded from the primary analysis because they were used to derive the corresponding omics scores. Including measured SBP and DBP as covariates would introduce potential redundancy and obscure the independent predictive contribution of the omics scores.

The MESA dataset was randomly partitioned into training (80%) and internal test (20%) sets, where training set was used to select hyperparameters via a grid search with 5-fold cross-validation, optimizing for held-out AUC. The tuning grid spanned the number of boosting rounds (100, 200, 300), maximum tree depth (3, 4, 6), learning rate (0.01 to 0.20 across 10 evenly spaced values), and minimum child node weight (1, 5, 8, 10). Column subsampling fraction and row subsampling fraction were fixed at 0.8 to provide regularization. Optimal hyperparameters identified were used to train the final model using the full (100%) MESA dataset for each prediction model to assess generalizability and transfer learning approaches to the JHS population.

#### Transfer learning

For transfer learning, JHS participants were randomly partitioned into an adaptation set (30-50%) and a held-out test set. The adaptation set was used to update MESA-trained models using either continued training or leaf refresh, whereas the held-out test set was reserved exclusively for external evaluation. The first strategy, continued training, extended the gradient boosting process by training additional rounds on JHS adaptation data. The optimal number of additional boosting rounds was determined by 3-fold cross-validation on each adaptation set, with the median selected across folds to reduce cross-fold variability. The second strategy, leaf refresh preserved the MESA-trained tree structure while refitting only the terminal leaf weights using JHS adaptation data. This approach recalibrated predicted probabilities to reflect outcome prevalence and feature distributions in JHS while preserving the biological relationships encoded in the original tree structure.

#### Model evaluation

Model performance was evaluated in the 20% MESA test set (when developed using 80% of MESA as training dataset), and independent external validation on full JHS data (when developed using 100% of MESA as the training dataset), across two complementary criteria, discrimination and calibration. Discrimination was quantified by the area under the receiver operating characteristic curve (AUC), while calibration was assessed by fitting a logistic model regressing the observed binary outcomes on the logit-transformed predicted probabilities, from which the calibration intercept and slope were derived. Validation datasets (either 20% MESA in the MESA-specific analysis or part of JHS in the second) were withheld from all model training and hyperparameter tuning procedures and used exclusively for unbiased estimation. All performance metrics were reported with 95% bootstrap confidence intervals (CI) derived from 1,000 bootstrap resamples. To enhance model interpretability, SHAP values were computed using R package shapviz (v0.10.1)^40^ for predictions in both the MESA-specific internal and JHS external validation data to characterize the contributions of individual features. For each transfer learning method, the best-performing model across the three adaptation sets was selected based on both discrimination and calibration in the held-out JHS validation set.

We conducted three sensitivity analyses. First, we refit all five models from the primary analysis including SBP and DBP as additional predictors to evaluate model performance when direct BP measurements are available. Moreover, to assess early prediction capability among individuals at low clinical risk, all fitted models were tested on JHS subgroup with normal baseline BP (SBP 100-120 mmHg and DBP 60-80 mmHg), excluding participants with elevated BP below the hypertension threshold at baseline. In addition, we developed MESA-specific ProtS and MetS using 80% of the MESA participants and used these scores to fit the corresponding XGBoost models, which were evaluated in the remaining 20% validation set. This analysis was performed to assess model performance in a similar population to that in which the scores were developed and to provide a reference for comparison with external validation in the JHS population.

#### Biomarker prioritization

To identify proteins with potential causal effects on BP traits and hypertension, we performed two-sample MR on proteins selected by Elastic Net using the TwoSampleMR R package (v0.7.5)^41^. This analysis aimed to distinguish proteins with potential causal effects on BP regulation from those whose predictive associations may reflect confounding or reverse causation. The Wald ratio method^42^ was applied to proteins with a single genetic instrument, and inverse-variance weighted (IVW) MR was used for proteins with multiple independent instruments.

Protein quantitative trait loci (pQTL) summary statistics from the UK Biobank^43^ were used as the exposure dataset, with genetic instruments restricted to cis-acting variants within ±1 Mb flanking window. For African ancestry analyses, adaptive posterior mean (APM)^44^ method was applied to improve protein instrument availability by improving African ancestry protein GWAS results. The APM method, applied for each available genetic variant association, borrows information from the European ancestry protein genetic association into the corresponding African ancestry association estimate in an adaptive manner, depending on the similarity of the estimates between European and African ancestry genetic associations. Outcome summary statistics were obtained from the MVP GWAS^45^ for four BP traits: SBP, DBP, hypertension, and essential hypertension, specific to EUR and AFR HARE (Harmonizing genetic ancestry and Race/Ethnicity) groups. These were matched in MR to the European and African ancestry protein genetic associations.

Genetic instruments were selected at genome-wide significance (p < 5×10^−8^) and filtered to single nucleotide polymorphisms (SNPs), excluding insertions, deletions, and variants on sex chromosomes to facilitate reliable allele harmonization. Instruments were clumped using PLINK^33^ (r^2^ < 0.001, 10,000-kb window) with ancestry-matched 1000 Genomes Project reference panels^46^ for European (EUR) and African (AFR) ancestries separately. To maximize the number of informative instruments per protein, only variants present in the outcome GWAS were retained prior to LD clumping. Instrument strength was assessed using the F-statistic. Protein associations with false discovery rate (FDR) < 0.05 were considered statistically significant within each BP outcome. All analyses were performed separately in EUR and AFR ancestry groups.

Bayesian colocalization analysis was performed for all proteins with significant MR associations using the R package coloc (v5.2.3) to determine whether the pQTL and BP GWAS association signals were driven by a shared causal variant. For each protein-outcome pair, pQTL and BP GWAS summary statistics were extracted for the corresponding genomic region and harmonized by chromosomal position. Duplicate variants were resolved by retaining the variant with the smallest P value. Significant colocalization was defined as a posterior probability of a shared causal variant (PP.H4) greater than 0.80.

#### Enrichment analysis

MR significant proteins across BP outcomes and ancestry groups were submitted to the STRING database^47^ (version 11.5) to construct a protein–protein interaction network. Functional enrichment analysis was performed for the resulting protein-network clusters using the R package enrichR^48,49^, querying for biological pathways annotated in Gene Ontology Biological Process and KEGG databases.

To assess the translational relevance of the identified proteins, we mapped the MR significant proteins to the Open Targets database^50^ to characterize existing evidence for cardiovascular disease and therapeutic tractability. The disease terms matching hypertension, blood pressure, cardiovascular, cardiac, coronary, heart, and arterial conditions were used to derive a cardiovascular association score (0-1). As a result, we classified the proteins into four categories: 1) established targets: proteins with well-established cardiovascular associations and existing therapeutic targets; 2) novel drug targets: proteins with cardiovascular evidence and drug tractability but no approved drug agents, representing promising opportunities for potential therapeutic development; 3) repurposing candidates: proteins with approved drugs but no established cardiovascular or BP effects; 4) novel findings: genetically validated proteins lacking prior evidence for cardiovascular diseases or therapeutic targeting, thereby warranting functional follow-up investigation.

## Results

Baseline characteristics for MESA and JHS participants stratified by hypertension status are summarized in Table 1. Overall, MESA participants were older and had a lower mean body mass index (BMI) than JHS participants across both hypertension subgroups (Table 1). Lifestyle factors also differed between cohorts, with higher proportions of current smoking status and alcohol use in MESA than in JHS (Table 1). Hypertension prevalence was lower in MESA (44.6%) than in JHS (61.5%), with hypertensive participants being older and having higher BMI than non-hypertensive participants in both cohorts (Table 1). Moreover, Black participants accounted for 35% of hypertensive cases despite comprising only 26.5% of the overall MESA participants, consistent with the disproportionate burden of hypertension among Black Americans. Among participants without hypertension at baseline, which constituted the population used for incident hypertension prediction, both mean SBP and DBP were higher in JHS (SBP: 120.4 ± 11.5, DBP: 75.0 ± 7.6 mmHg) than in MESA (SBP: 115.7 ± 13.4, DBP: 69.3 ± 8.8 mmHg). These baseline differences in hypertension prevalence and BP distributions between MESA and JHS have direct implications for model generalizability and motivates the transfer learning approach and sensitivity analysis employed in the study.

**Table 1.** Baseline Characteristics of Study Participants by Baseline Hypertension (HTN) Status in MESA and JHS.

|  | MESA |  |  | JHS |  |  |
|---|---|---|---|---|---|---|
| **Characteristic** | **Overall**<br>(N = 6290)<sup>1</sup> | **No HTN**<br>(N = 3486)<sup>1</sup> | **HTN**<br>(N = 2804)<sup>1</sup> | **Overall**<br>(N = 3160)<sup>1</sup> | **No HTN**<br>(N = 1215)<sup>1</sup> | **HTN**<br>(N = 1945)<sup>1</sup> |
| **Age** (years) | 62.23 (10.25) | 59.32 (9.88) | 65.86 (9.51) | 56.40 (12.44) | 50.14 (12.09) | 60.31 (10.96) |
| **Sex** |  |  |  |  |  |  |
| **Female** | 3,299.0 (52.4%) | 1,766.0 (50.7%) | 1,533.0 (54.7%) | 2,018.0 (63.9%) | 746.0 (61.4%) | 1,272.0 (65.4%) |
| **Male** | 2,991.0 (47.6%) | 1,720.0 (49.3%) | 1,271.0 (45.3%) | 1,142.0 (36.1%) | 469.0 (38.6%) | 673.0 (34.6%) |
| **Race/Ethnicity** |  |  |  |  |  |  |
| **White** | 2,476.0 (39.4%) | 1,520.0 (43.6%) | 956.0 (34.1%) | 0.0 (0.0%) | 0.0 (0.0%) | 0.0 (0.0%) |
| **Chinese-American** | 712.0 (11.3%) | 446.0 (12.8%) | 266.0 (9.5%) | 0.0 (0.0%) | 0.0 (0.0%) | 0.0 (0.0%) |
| **Black** | 1,664.0 (26.5%) | 683.0 (19.6%) | 981.0 (35.0%) | 3,160.0 (100.0%) | 1,215.0 (100.0%) | 1,945.0 (100.0%) |
| **Hispanic** | 1,438.0 (22.9%) | 837.0 (24.0%) | 601.0 (21.4%) | 0.0 (0.0%) | 0.0 (0.0%) | 0.0 (0.0%) |
| **BMI** (kg/m²) | 28.35 (5.48) | 27.48 (5.18) | 29.43 (5.65) | 32.03 (7.32) | 30.98 (7.37) | 32.69 (7.21) |
| **Smoking Status** |  |  |  |  |  |  |
| **Never/Former** | 3,152.0 (57.9%) | 1,731.0 (58.6%) | 1,421.0 (57.1%) | 2,127.0 (77.1%) | 874.0 (83.6%) | 1,253.0 (73.1%) |
| **Current** | 2,292.0 (42.1%) | 1,223.0 (41.4%) | 1,069.0 (42.9%) | 631.0 (22.9%) | 171.0 (16.4%) | 460.0 (26.9%) |
| **Alcohol Use** | 3,463.0 (68.6%) | 2,063.0 (72.0%) | 1,400.0 (64.0%) | 1,435.0 (45.6%) | 670.0 (55.4%) | 765.0 (39.5%) |
| **Time Between Visits** (years) | 9.47 (0.65) | 9.50 (0.65) | N/A | 7.49 (1.40) | 7.49 (1.40) | N/A |
| **SBP** (mmHg) | 132.09 (24.68) | 115.71 (13.44) | 152.44 (19.85) | 136.68 (19.56) | 120.43 (11.49) | 146.89 (16.43) |
| **DBP** (mmHg) | 75.52 (11.84) | 69.29 (8.83) | 83.27 (10.44) | 81.59 (10.00) | 75.00 (7.62) | 85.72 (9.06) |

<sup>1</sup>Mean (SD); n (%). Time between visits was calculated for participants without hypertension at baseline and is not applicable (N/A) to those with prevalent hypertension.

### Metabolite and protein scores for BP traits

The metabolites and proteins selected for score construction are listed in the Supplementary Data, along with their corresponding model coefficients. We examined associations of BP related ProtS, MetS, and PRS with corresponding BP traits in all available JHS participants (Table S2). All three score types were significantly associated with their corresponding phenotypes. ProtS demonstrated the strongest associations across all BP traits, explaining 16.1% and 17.8% of variance in SBP and DBP respectively (SBP: β=0.72, p=1.96×10^−46^; DBP: β=0.82, p=2.36×10^−43^), and an odds ratio of 2.05 for HTN (p=2.50×10^−21^). In contrast, MetS and PRS showed significant but substantially attenuated associations with corresponding BP traits, explaining markedly less variance than ProtS despite a larger available sample (Table S2). These results confirm that omics scores derived in MESA retain biological relevance and portability in JHS.

### XGBoost models for incident hypertension

The optimal hyperparameters identified through 5-fold cross-validation for the primary and sensitivity analyses are presented in Table S3. In the primary analysis, all models converged to shallow trees (max_depth = 3), indicating that hypertension prediction based on omics scores can be adequately captured by relatively simple decision boundaries without requiring higher-order feature interactions. The MetS model required the largest number of boosting rounds (nrounds = 300), suggesting weaker predictive signal from individual MetS relative to ProtS and PRS (Table S3). In the sensitivity analysis adding SBP and DBP as model features, the ProtS and all-scores models selected a maximum tree depth of 4, indicating additional complex interactions introduced by measured BP (Table S3). Sensitivity analyses using scores developed on 80% of MESA selected a similar hyperparameter profile to the primary analysis, with the exception of min_child_weight (Table S3).

### Model evaluation and transfer learning

In the MESA-specific analysis using the 20% held-out test data, model discrimination increased progressively with the addition of omics scores, with the AUC increasing from 0.63 for the baseline clinical model to 0.72 for the combined all-scores model (Figure 2A). In the analysis with JHS external validation (with models developed using the complete MESA dataset), all models showed lower AUCs than in the MESA-specific evaluation, reflecting the cross-cohort generalization gap. Notably, the ProtS model achieved the highest AUC of 0.69 in JHS and the smallest decline in performance compared to that in MESA test set (Figure 2A), indicating superior discrimination and transferability across cohorts. The large gap observed between MESA (AUC= 0.72) and JHS (AUC= 0.65) all-scores model may suggest overfitting to MESA-specific biological patterns when all omics layers are combined.

**Figure 2.**
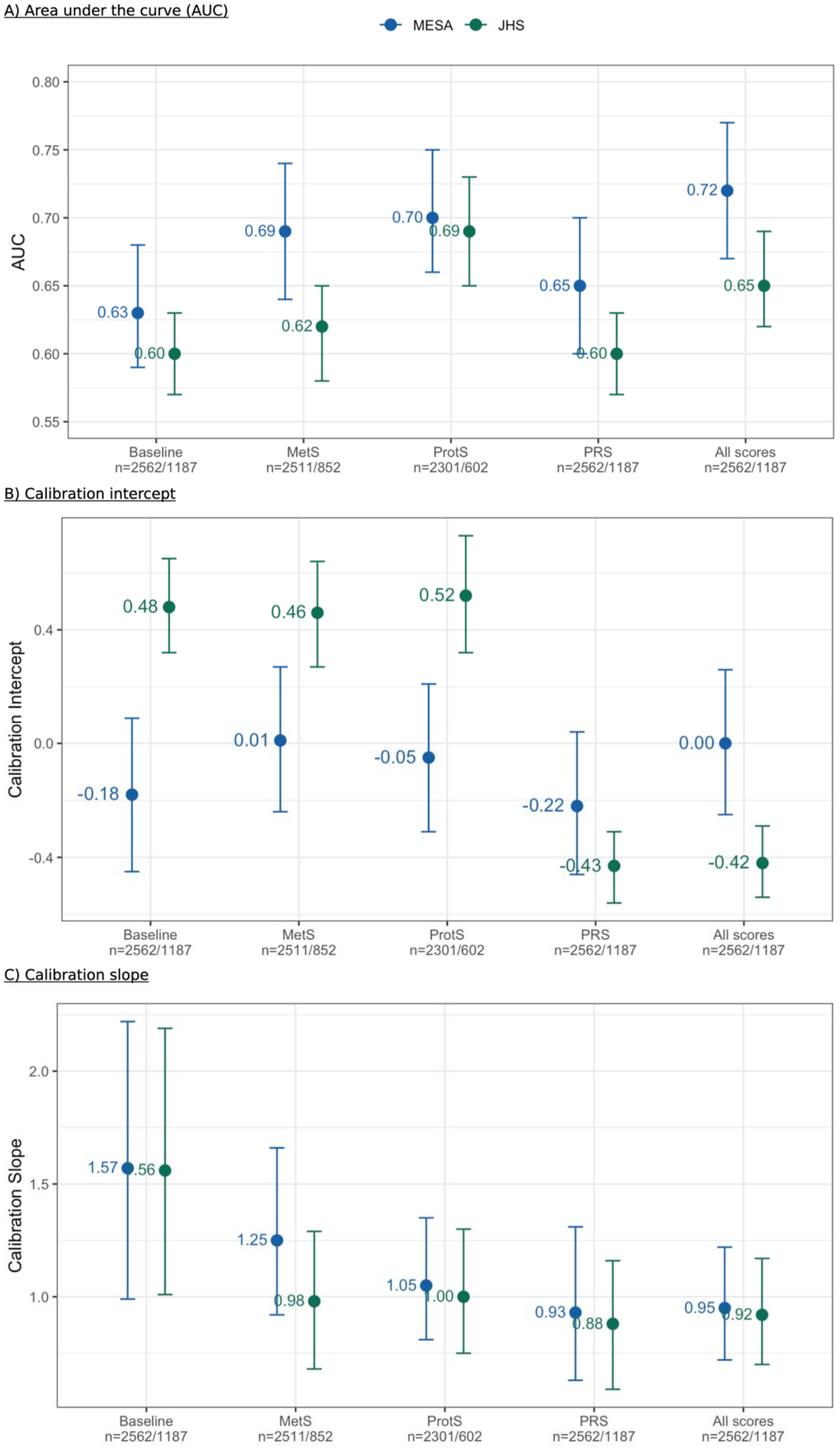
Discrimination and calibration of multi-omics XGBoost models for incident hypertension prediction across MESA cohort-specific evaluation and JHS cohort-generalized evaluation in primary analysis. A) Area under the curve (AUC), B) calibration intercept, and C) calibration slope for five progressively richer models: baseline clinical covariates (age, sex, self-reported race, BMI, smoking status, alcohol use, and time between study visits), metabolomic scores (MetS), proteomic scores (ProtS), polygenic risk scores (PRS), and all omics scores combined. Models were evaluated on the held-out 20% MESA test set (blue; cohort-specific analysis), and on the full JHS cohort (green; cohort-generalized analysis). Points represent point estimates and error bars represent 95% bootstrap confidence intervals. Sample sizes for each model are reported as n= n(MESA)/n(JHS).

Calibration performance is presented in Figure 2B–C. All cohort-specific models were generally well calibrated, with calibration intercepts and slopes whose 95% CI included the ideal values of 0 and 1 respectively. In contrast, calibration patterns differed across models in cohort-generalized analysis (Figure 2B-C). Specifically, the baseline, MetS, and ProtS models underpredicted hypertension risks, as indicated by positive calibration intercepts, in comparison to overprediction in the PRS and all-scores models (Figure 2B). These opposing calibration patterns across model types motivate the application of transfer learning to recalibrate predictions in JHS. Calibration slopes also varied across models (Figure 2C), with the baseline clinical model exhibiting the greatest deviation from the ideal value of 1 and the widest CI in both MESA (1.57) and JHS (1.56). The addition of omic scores greatly improved calibration, with slopes shifting closer towards 1 (Figure 2C).

To address the calibration gap observed between MESA and JHS, we applied two transfer learning strategies, continued training and leaf weight refresh. While both methods produced little to modest improvement in discrimination across the five models (Figure 3A), leaf refreshing achieved substantial improvements in brining calibration intercept towards 0, despite a wider CI (Figure 3B). Similarly, leaf refreshing achieved calibration slopes comparable to those obtained with continued training but with greater uncertainty, as reflected by wider CI (Figure 3C). The wider CIs observed for the two transfer learning methods may be explained, at least to some degree, by the smaller sample sizes left for evaluation, as a subset of JHS data was used for transfer learning, leading to more variable prediction performance estimates. Among the 5 models, the baseline clinical model demonstrated the greatest instability in calibration estimation and adaptation (Figure 2-3). Overall, leaf refresh stood out as the preferred transfer learning strategy, providing the greatest improvements in calibration while maintaining comparable discrimination performance. This indicates that the predictor–outcome relationships largely generalize to JHS, whereas the terminal leaf predictions require recalibration to account for cohort-specific characteristics.

**Figure 3.**
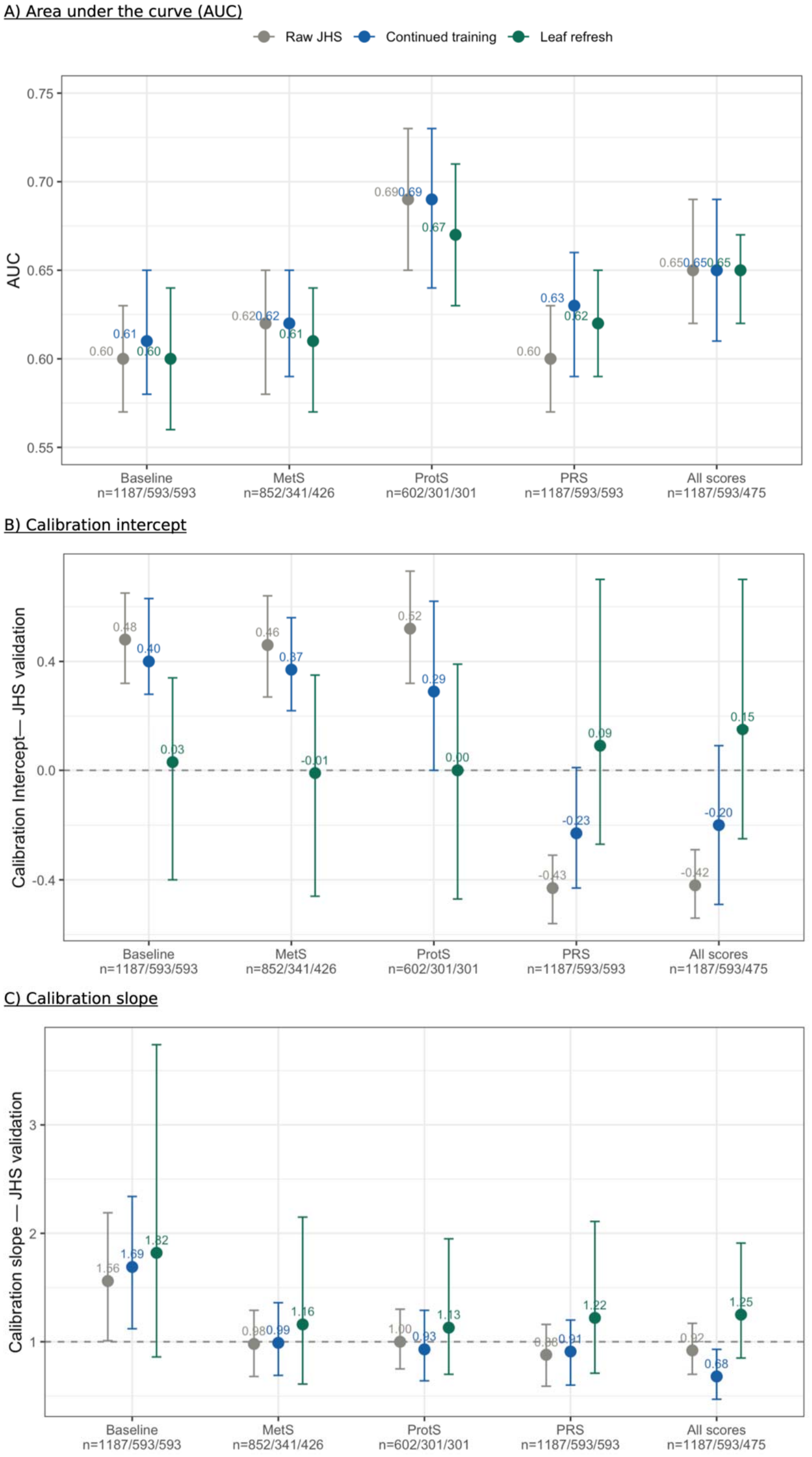
Effect of transfer learning strategies on XGBoost models in JHS external validation. A) Area under the curve (AUC), B) calibration intercept, and C) calibration slope in JHS external validation for raw model without adaptation (grey), continued training (blue), and leaf weight refresh (green). Dashed lines indicate ideal calibration (intercept = 0, slope = 1). Sample sizes for each model are reported as n= n(raw JHS evaluation)/n(continued training)/n(leaf refreshing).

### Sensitivity analysis and early prediction in JHS normal BP group

To evaluate the ability of omics scores to provide predictive value independent of BP measurements, we repeated our primary analyses including SBP and DBP as model predictors. The optimal hyperparameters for these BP-inclusive models are also reported in Table S3. Compared to the primary analysis, ProtS and all-scores models required greater complexity and more boosting rounds (Table S3). Interestingly, the PRS+BP model retained identical hyperparameters as in primary analysis, suggesting that PRS and BP measurements contribute relatively independent additive signals. The inclusion of measured SBP and DBP boosted discrimination across all models, with narrower generalization gap (Figure S1A). In cohort-specific models evaluations dataset, AUC ranged from 0.76 to 0.80 for PRS+BP model (Figure S1A). In the cohort-generalized evaluation dataset, the simplest baseline+BP model achieved an AUC of 0.74, while ProtS+BP model reached the highest AUC of 0.75 (Figure S1A), indicating that ProtS provided additional discrimination power in the presence of BP readings. Calibration metrics showed similar patterns to those observed in primary analysis (Figure S1B-C), confirming that the miscalibration is a cross-cohort phenomenon.

Figure S2 and S3 summarize mean absolute SHAP values for all five prediction models in primary and sensitivity analyses, respectively. Across both MESA and JHS, ProtS_SBP was the dominant feature in ProtS-only model (MESA: 0.344; JHS: 0.337), but such importance was attenuated in all-scores model, particularly in JHS (MESA: 0.3; JHS:0.189) (Figure S2). Age and BMI ranked among the most influential predictors across all models in both cohort-specific and cohort-generalized analyses, consistent with their well-established roles as major hypertension risk factors. Notably, BMI contributed more strongly to model predictions in JHS than in MESA across all fitted models (Figure S2-S3). Compared to ProtS and PRS, MetS exhibited the largest reduction in SHAP importance when other type of scores or direct BP measurements were incorporated, especially in JHS (Figure S2-S3). Race-AFR showed substantially higher importance in JHS; this is likely due to all participants in JHS having the same value for this term, unlike in MESA, as all JHS participants self-identify as Black or African American based on the study design. The Race-AFR term becomes less important in the PRS and all-scores models in primary analysis (Figure S2) in JHS due to adjustment for genetic PCs, which capture some of the variance attributed to the Race-AFR term in other models.

When measured SBP and DBP were included as model features, SBP emerged as the dominant predictor across all models in both cohorts, whereas DBP contributed modestly (Figure S3). Still, ProtS_SBP retained meaningful importance in sensitivity analysis (Figure S3), corroborating that ProtS add independent biological signal beyond contemporaneous BP status for incident hypertension prediction. Demographic predictors with large SHAP values in primary analysis, such as age, BMI, and Race-AFR, experienced substantial reduction in BP inclusive models (Figure S2-S3). Moreover, SBP attained its highest SHAP value in the PRS+BP model, suggesting that genetic predisposition may enhance how the model utilizes BP information (Figure S3).

To evaluate the potential for early hypertension prediction before individuals meet clinical blood pressure criteria for elevated risk, we applied all fitted models to the subset of JHS participants with normal baseline BP (SBP 100-120 mmHg, DBP 60-80 mmHg). Same as primary analysis, the ProtS model showed the strongest discrimination among models without SBP and DBP, with the highest AUC of 0.63, same as the baseline+BP model (Figure 4). However, it also showed the widest CI, likely reflecting the limited sample size of this subgroup (n= 167). The consistent performance of ProtS across cohorts and blood pressure strata indicates that circulating proteins capture hypertension-relevant biological signals which can generalize acros populations and provide predictive power before clinically detectable BP elevations. The combination of omic scores and measured SBP and DBP improved discrimination relative to baseline models, especially the ones including PRS, with all-scores+BP model reaching an AUC of 0.7 (Figure 4), suggesting that constitutional genetic risk provides complementary predictive information beyond measured BP, particularly for identifying individuals at increased risk of developing hypertension despite having clinically normal blood pressure at baseline.

**Figure 4.**
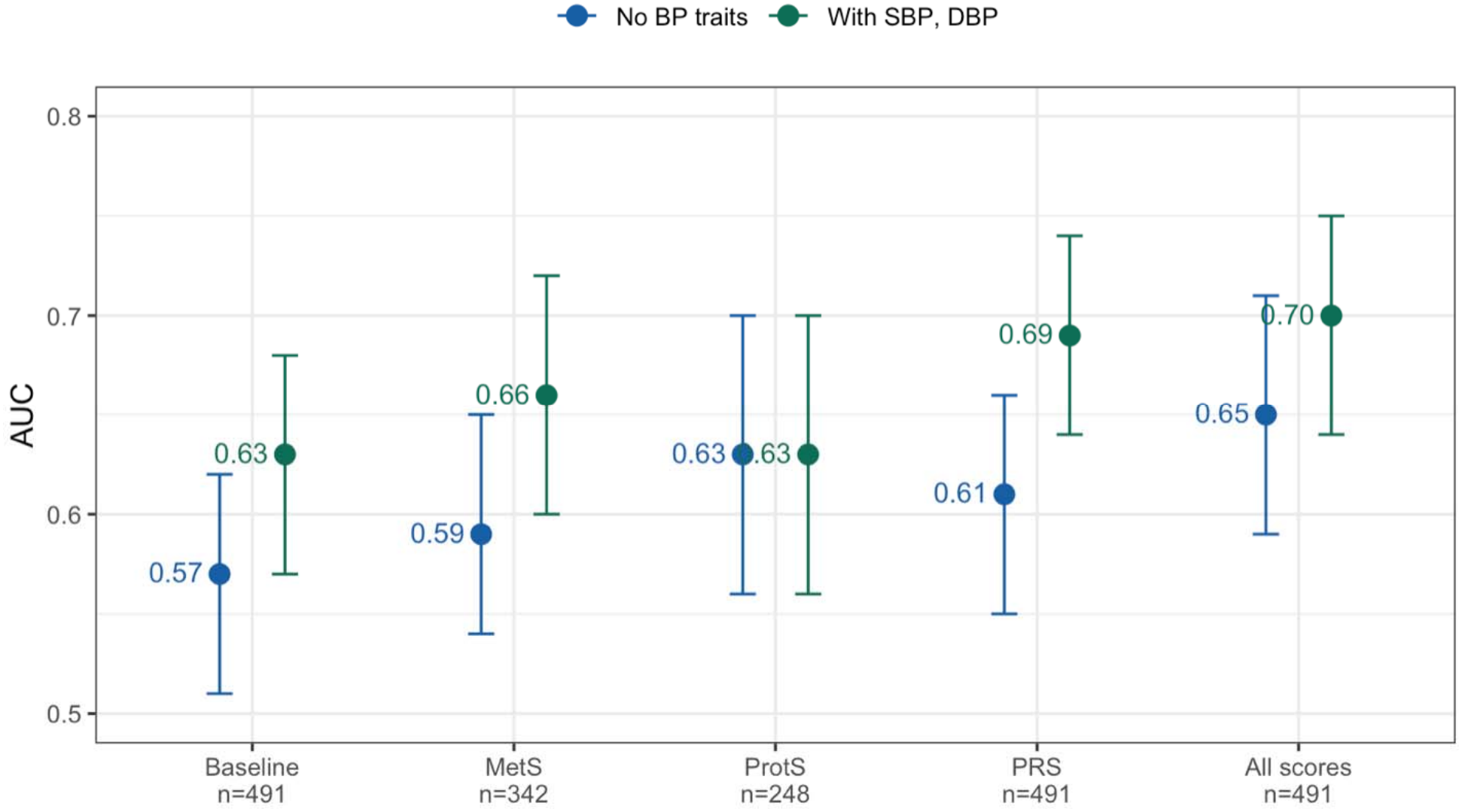
Early prediction of incident hypertension in JHS participants with normal blood pressure (SBP 100-120 mmHg, DBP 60-80 mmHg) at baseline. Sample sizes for each model are reported as n.

Models using ProtS and MetS derived from 80% of MESA participants yielded comparable and consistent performance with models using omics scores from the full MESA cohort, with overlapping 95%CI (Table S4). These results indicate that both score derivation and model performance were robust to moderate differences in the sample size used for Elastic Net.

### Biomarker prioritization

In the European ancestry, MR identified 29 proteins causally associated with SBP, 34 with DBP, and 19 with hypertension or essential HTN at an FDR < 0.05 (Supplementary Data 2). All genetic instruments had F-statistics exceeding the conventional threshold of 10 (Supplementary Data 2), confirming strong instrument strength. The majority of the MR significant proteins were identified using the Wald ratio method, reflecting that most proteins had a single genome-wide significant instrument after merging with outcome GWAS and LD clumping. We also identified 5 MR proteins for SBP, and 1 for both DBP and hypertension in the African ancestry analysis, out of which inositol-requiring enzyme 1 alpha (ERN1) for SBP, ectonucleoside triphosphate diphosphohydrolase 5 (ENTPD5) for DBP, and N-terminal pro-B-type natriuretic peptide (NTproBNP) for hypertension were only discovered in this group (Supplementary Data 2).

In EUR ancestry, four MR significant proteins demonstrated strong colocalization evidence (PP.H4> 0.80), spanning seven independent genomic loci (Table 2). The colocalization results and limited MR proteins identified for African ancestry likely reflected the limited instrument availability and statistical power due to smaller sample sizes and correspondingly sparser variant coverage in the tested genomic regions.

**Table 2.** Colocalization results in EUR ancestry analysis.

| **Exposure** | **SNP** | **CHR** | **Position** | **Outcome** |
|---|---|---|---|---|
| FGF5 | rs6821485 | 4 | 80780882 | SBP |
| CCL28 | rs1593971 | 5 | 43413342 | DBP |
| WARS | rs2273804 | 14 | 100376300 | SBP |
| WARS | rs1555405 | 14 | 100710432 | SBP |
| AGT | rs2478539 | 1 | 230709026 | HTN |
| AGT | rs7540252 | 1 | 230746703 | HTN |
| AGT | rs34943600 | 1 | 230915795 | HTN |

CHR: chromosome; FGF5: fibroblast growth factor 5; CCL28: C-C motif chemokine ligand 28; WARS: tryptophanyl-tRNA Synthetase 1; AGT: angiotensinogen. SBP, DBP: systolic and diastolic blood pressure; HTN: hypertension.

### Enrichment analysis

Protein–protein interaction analysis of MR-significant proteins (identified in either EUR or AFR analyses) using the STRING database identified a densely connected central network, consolidating into three tightly interconnected modules, cross-linked by Serpin Family E Member 1 (SERPINE1), endothelin 1 (EDN1), and interleukin-8 (CXCL8) (Figure S5). The first cluster can be described as the coagulation and vascular integrity module (cluster 1), represented by SERPINE1, cadherin-5 (CDH5), annexin A2 (ANXA2), and tryptophanyl-tRNA Synthetase 1 (WARS) (Figure S5). The Renin-Angiotensin-Aldosterone System (RAAS) and natriuretic signaling module (cluster 8) was characterized by angiotensinogen (AGT), endothelin 1 (EDN1), and two forms of natriuretic peptides B (NPPB, NTproBNP) (Fiure S4). The inflammatory and extracellular matrix remodeling module (cluster 2) involved interleukin-8 (CXCL8), transforming growth factor beta-1 proprotein (TGFB1), and aggrecanase-1 (ADAMTS4). Functional enrichment analysis demonstrated that this central network was significantly enriched for biological pathways implicated in hypertension pathogenesis, including TGF-β receptor signaling, regulation of inflammatory response, plasminogen activation, vasculature development and renal sodium excretion (Figure S5). Collectively, these results supported a causal cascade towards HTN involving coherent inflammation, renal sodium handling and vascular remodeling.

Among the 77 MR significant proteins, 11 were classified as established targets with both cardiovascular disease evidence and existing approved or investigational therapeutics, such as AGT, a well-established therapeutic target currently under clinical investigation, and proprotein convertase subtilisin/kexin type 9 (PCSK9), which showed the highest cardiovascular evidence score of 0.72 (Table S5). 29 proteins were classified as novel drug targets, among which NPPB was statistically significant with SBP, and hypertension in both European and African groups, making it a high-potential drug target given its strong causal evidence and antibody tractability (Table S5). While majority of the MR proteins showed cardiovascular evidence, 34 were identified as potential novel findings, such as WARS, a colocalized protein showing causal associations with both SBP and DBP in European ancestry (Table S5).

## Discussion

This study presents a systematic evaluation of multi-omics scores for incident hypertension prediction with external validation in an independent cohort. Our results demonstrate that omics scores improve hypertension risk prediction beyond clinical covariates, with ProtS consistently showing strong predictive performance and cross-cohort transferability. This finding aligns with the biological nature of distinct omics layers^51,52^. Compared to proteomics, metabolomic profiles are more sensitive to variation^53^ arising from dietary habits, environmental exposures, and lifestyle factors, which likely differ between MESA and JHS. In addition, the substantially smaller number of annotated metabolites available in JHS may have further contributed to reduced transferability of MetS. Consequently, MetS explained less variance of SBP and DBP, and exhibited weaker predictive performance and larger generalization gap.

Our findings also reaffirm the robustness of SBP and DBP as the strongest clinical predictors of incident hypertension^54^ . As models incorporating these BP measurements achieved the highest AUC in both overall study population and the subgroup with normal BP. Although this result is expected given that the omic scores were derived using these BP traits, ProtS_SBP retained meaningful SHAP importance even when measured SBP was included as a predictor (Figure S2). Consistent with a recent study^55^, our results corroborated that ProtS capture biological signal of hypertension risk independent of BP measurements.

Among the relatively few studies incorporating omics data in HTN prediction^55–58^, PRS have consistently provided greater improvements in incident hypertension prediction than metabolomic, transcriptomic, or gut microbiome data when added to clinical risk factors, although the discrimination gain is rather modest than transformative. In alignment with previous research, our PRS+BP model also demonstrated best discrimination (AUC=0.8) in MESA internal validation, but only a marginal improvement over the corresponding baseline + BP model (Figure S1). Another particularly noteworthy finding was observed in the JHS subgroup with normal baseline BP, where PRS+BP model and the ProtS model without SBP and DBP demonstrated superior performance, providing evidence for PRS and circulating proteins in capturing biological predisposition prior to detectable BP elevation. This clinically impactful finding is of special interests for individuals whose BP measurements at a single timepoint may underestimate their true long-term hypertenison risk. Together, these findings warrant future investigation of ProtS and PRS in identifying high-risk individuals within a window in which preventive interventions may be most effective but conventional clinical assessment remains insufficient.

The observed generalization gap is likely attributable to the heterogeneity between MESA and JHS, spanning from genetic ancestry differences, environmental and lifestyle factors, to markedly higher hypertension prevalence in JHS. Moreover, SBP and DBP inclusive models showed better discrimination but worsened calibration slopes than models based solely on omics scores (Figure 2 and S1). This trade-off indicates that the risk ranking may be pushed by extreme BP values, which generalize less well across populations. In contrast, a relatively more robust transferability observed for omic scores suggests that they capture more stable biological processes underlying hypertension risk. Given that BP is inherently labile and can fluctuate in response to medications, stress, and even without overt provocations^59^, our results may further indicate a stronger resistance of omic scores to BP variability. Consistent with this interpretation, the leaf refresh transfer learning strategy for gradient boosted decision trees was particularly effective at correcting calibration while preserving discrimination. As in this case, miscalibration arose primarily from differences in disease prevalence and cohort characteristics rather than structural model misspecification.

MR findings provide mechanistic insight into the circulating proteins, collectively describing the causal cascade of BP regulation. As the sole precursor of the RAAS pathway, genetically predicted higher circulating AGT was causally associated with increased hypertension risk, likely through greater substrate availability for angiotensin II generation, thereby promoting vasoconstriction^60^. Conversely, brain natriuretic peptide (NPPB) and its co-secreted inactive form NTproBNP exhibited the strongest protective effects against hypertension in both European and African ancestry groups (Table S5). In response to BP overload, ventricular myocytes release NPPB to enhance natriuresis, vasodilation, and suppression of RAAS cascade^61^, consistent with our MR findings supporting a protective role of genetically predicted NPPB/NT-proBNP on hypertension risk (Table S5).

Given that elevated circulating NTproBNP levels in people with high BP were associated with increased cardiovascular mortality, the substantially higher protein level in both MESA (Welch’s t-test, mean difference= 0.415, p < 2.2e-16) and JHS hypertension group (mean difference= 0.437, p = 4.185e-12) further support NTproBNP as a potential biomarker of greater cardiovascular burden among individuals with hypertension, such as coronary artery disease, heart failure, and atrial fibrillation^18^. Despite this robust genetic evidence, no therapies currently target NPPB directly for hypertension, although neprilysin inhibitors and angiotensin receptor blockers enhance natriuretic peptide signaling and are widely used together in heart failure treatment, with additional antihypertensive benefits ^62^.

EDN1, one of the most potent endogenous vasoconstrictors emerged as a central hub protein in the STRING network and demonstrated a causal risk-increasing effect on SBP. Along with TGFB1, CDH5, CXCL8, and SERPINE1, these central hub proteins were enriched in positive regulation of vasculature development. Therefore, altered circulating concentrations of these proteins may reflect subclinical BP dysregulation preceding overt hypertension, through mechanisms involving arterial fibrosis, endothelial dysfunction, vascular smooth muscle stiffening, and chronic low-grade inflammation^63–66^.

Colocalization evidence supported that two novel variants, rs2273804 and rs1555405, of WARS (or WARS1) and rs1593971 of C-C motif chemokine ligand 28 (CCL28), both implicated in inflammatory pathways, can causally raise SBP and DBP levels, respectively (Table 2). Another WARS-associated variant rs724391 has previously been linked to greater hypertension risk^67^, potentially through increased peripheral vascular resistance mediated by endothelial permeability^68^ and inflammation^69^. On the other hand, CCL28 is a mucosal-homing chemokine implicated in an innate immune defense^70^, and has been shown in mouse models to promote cardiac repair by stimulating angiogenesis^71^. Accordingly, our findings suggest that WARS and CCL28 may represent novel mechanistic links between BP dysregulation and hypertension-related end-organ damage, possibly via systemic hyperinflammation and mucosal immune signaling.

Strengths of the study include the systematic comparison of ProtS, MetS, and PRS contributions to hypertension prediction within a unified framework, allowing assessment of both predictive performance and cross-cohort generalizability. We further validated the superior predictive performance of clinical blood pressure measurements for incident hypertension. This study also presents several limitations. First, the number of JHS participants with available proteomic data was relatively small (N= 398), with only 167 remaining in the subgroup with normal baseline BP. This limited sample size likely contributed to the wide CI observed for ProtS model. Second, due to the nature of cross-sectional cohorts, exact time of hypertension diagnosis and information on hypertension subtypes were unavailable. The higher baseline BMI and greater SHAP importance of BMI in JHS suggest that obesity-driven hypertension may contribute more to disease risk in this cohort, meanwhile exhibiting distinct molecular signatures. Previous studies have shown promising results using proteomics and multi-omics panels to distinguish primary aldosteronism and endocrine subtypes from primary hypertension^72–74^, highlighting the opportunity for future work towards hypertension subtype classification and mechanism-guided precision medicine. In addition, the omics scores were derived in MESA and thus may carry cohort specific biological patterns, which could lead to marginally optimistic internal validation estimates. Although external validation in JHS supports generalizability of the findings, replication in larger and more diverse cohorts is warranted, particularly among individuals with normal BP. Finally, clinical implementation of proteomic data requires continued technological advances including cost reduction, harmonization of profiling assays and broader access to high-throughput protein technologies.

## Conclusions

In conclusion, integration of omics scores with machine learning improved prediction of incident hypertension beyond clinical covariates, with ProtS showing the most consistent performance and cross-cohort transferability. Although measured SBP and DBP remained the strongest predictors of incident hypertension, ProtS retained predictive importance beyond baseline BP measurements, suggesting that circulating proteins capture biological signals of hypertension risk that are relatively preserved across cohorts. Combining BP and HTN PRS with BP measurements also showed potential for risk stratification among individuals without clinically elevated BP. Together, these findings support an integrated framework incorporating proteomic and genetic information for earlier and more precise hypertension risk stratification.

## Acknowledgement

Whole genome sequencing (WGS) for the Trans-Omics in Precision Medicine (TOPMed) program was supported by the National Heart, Lung and Blood Institute (NHLBI). WGS for “NHLBI TOPMed: Multi-Ethnic Study of Atherosclerosis (MESA)” (phs001416.v1.p1) was performed at the Broad Institute of MIT and Harvard (3U54HG003067-13S1). WGS for “NHLBI TOPMed: The Jackson Heart Study (JHS)” (phs000964.v6.p2) was performed at the Northwest Genomics Center (HHSN268201100037C). Centralized read mapping and genotype calling, along with variant quality metrics and filtering were provided by the TOPMed Informatics Research Center (3R01HL-117626-02S1). Phenotype harmonization, data management, sample-identity QC, and general study coordination, were provided by the TOPMed Data Coordinating Center (3R01HL-120393-02S1). The Multi-Ethnic Study of Atherosclerosis (MESA) projects are conducted and supported by the National Heart, Lung, and Blood Institute (NHLBI) in collaboration with MESA investigators. Support for MESA is provided by contracts 75N92025D00022, 75N92020D00001, HHSN268201500003I, N01-HC-95159, 75N92025D00026, 75N92020D00005, N01-HC-95160, 75N92020D00002, N01-HC-95161, 75N92025D00024, 75N92020D00003, N01-HC-95162, 75N92025D00027, 75N92020D00006, N01-HC-95163, 75N92025D00025, 75N92020D00004, N01-HC-95164, 75N92025D00028, 75N92020D00007, N01-HC-95165, N01-HC-95166, N01-HC-95167, N01-HC-95168, N01-HC-95169, UL1-TR-000040, UL1-TR-001079, UL1-TR-001420, UL1TR001881, and R01HL105756. The authors thank the MESA participants and the MESA investigators and staff for their valuable contributions. A full list of participating MESA investigators and institutions can be found at http://www.mesa-nhlbi.org

The Jackson Heart Study is supported by Contracts HHSN268201800010I, HHSN268201800011I, HHSN268201800012I, HHSN268201800013I, HHSN268201800014I, HHSN268201800015I from the National Heart, Lung, and Blood Institute (NHLBI) with additional support from the National Institute on Minority Health and Health Disparities (NIMHD).

The views expressed in this manuscript are those of the authors and do not necessarily represent the views of the National Heart, Lung, and Blood Institute; the National Institute of Minority Health and Health Disparities (NIMHD); the National Institutes of Health; or the U.S. Department of Health and Human Services.

UAT is supported by R01HL188729 and K08HL161445.

## Data Availability

TOPMed freeze 10 WGS, and phenotype data are available by application to dbGaP according to the study specific accession: JHS: “phs000964”, MESA: “phs001211”. MESA phenotype data are available from dbGaP according to the MESA accession: MESA: “phs000209”.

## Ethics approval and consent to participate

This work was approved by the Beth Israel Deaconess Medical Center Committee on Clinical Investigations, protocol #2023P000541. All MESA participants provided written informed consent, and the study was approved by the Institutional Review Boards at The Lundquist Institute (formerly Los Angeles BioMedical Research Institute) at Harbor-UCLA Medical Center, University of Washington, Wake Forest School of Medicine, Northwestern University, University of Minnesota, Columbia University, and Johns Hopkins University. The Institutional Review Boards at Jackson State University, Tougaloo College, and the University of Mississippi Medical Center approved the original study protocol, and all participants provided written informed consent. Ongoing IRB review for study activities is through the University of Mississippi Medical Center and Wake Forest University Health Sciences.

## Author contribution

Z.W conduct analysis, write and revise the manuscript; Y.Hrytsenko, S.Almogy process data, review and revise the manuscript; B.Spitzer compute polygenic risk scores; Y.Purushotham process data for mendelian randomization analysis; K.Taylor, J.Rotter, prepare and curate the MESA data; review and revise the manuscript; H.Lin, U.Tahir, Z.Chen curate data, review and revise the manuscript; L.Raffield prepare and curate the JHS data; review and revise the manuscript; T.Sofer conceive the study and supervise the analysis; review and revise the manuscript.

## References

1. Zhou, B., Perel, P., Mensah, G. A. & Ezzati, M. Global epidemiology, health burden and effective interventions for elevated blood pressure and hypertension. Nat. Rev. Cardiol. 18, 785–802 (2021).

2. NCD Risk Factor Collaboration (NCD-RisC). Worldwide trends in blood pressure from 1975 to 2015: a pooled analysis of 1479 population-based measurement studies with 19·1 million participants. Lancet 389, 37–55 (2017).

3. Flack, J. M., Ferdinand, K. C. & Nasser, S. A. Epidemiology of hypertension and cardiovascular disease in African Americans. J. Clin. Hypertens. (Greenwich) 5, 5–11 (2003).

4. Mann, S. J. The clinical spectrum of labile hypertension: a management dilemma. J. Clin. Hypertens. (Greenwich) 11, 491–7 (2009).

5. Kolifarhood, G. et al. Heritability of blood pressure traits in diverse populations: a systematic review and meta-analysis. J. Hum. Hypertens. 33, 775–785 (2019).

6. Padmanabhan, S. & Dominiczak, A. F. Genomics of hypertension: the road to precision medicine. Nat. Rev. Cardiol. 18, 235–250 (2021).

7. Fujii, R. et al. Associations of Genome-Wide Polygenic Risk Score and Risk Factors With Hypertension in a Japanese Population. Circ. Genom. Precis. Med. 15, (2022).

8. Kim, N. Y., Park, J., Nam, K., Lee, S. & Lee, H. Associations Between Polygenic Risk Score for Blood Pressure and Risk of Hypertension in Northeast Asian Individuals. J. Am. Heart Assoc. 15, (2026).

9. Kurniansyah, N. et al. Evaluating the use of blood pressure polygenic risk scores across race/ethnic background groups. Nat. Commun. 14, 3202 (2023).

10. Martin, A. R. et al. Clinical use of current polygenic risk scores may exacerbate health disparities. Nat. Genet. 51, 584–591 (2019).

11. Sun, C. et al. A review of disease risk prediction methods and applications in the omics era. Proteomics 24, (2024).

12. Menni, C. et al. Metabolomic Identification of a Novel Pathway of Blood Pressure Regulation Involving Hexadecanedioate. Hypertension 66, 422–429 (2015).

13. Holmes, E. et al. Human metabolic phenotype diversity and its association with diet and blood pressure. Nature 453, 396–400 (2008).

14. Liu, Y. et al. Identification of Blood Pressure-Associated Metabolites by Integrating Metabolomic and Genetic Analysis. MedComm (Beijing). 7, e70718 (2026).

15. Aldisi, R. S., Alsamman, A. M., Krawitz, P., Maj, C. & Zayed, H. Identification of novel proteomic biomarkers for hypertension: a targeted approach for precision medicine. Clin. Proteomics 22, 7 (2025).

16. Aggarwal, M. et al. Integrative Proteomic Profiling of Blood Pressure and Hypertension. Hypertension 83, (2026).

17. Du, J. et al. Plasma Biomarker Screening Based on Proteomic Signature of Patients with Resistant Hypertension. J. Cardiovasc. Transl. Res. 17, 1286–1294 (2024).

18. Luo, Y. et al. AI-based multiomics profiling reveals complementary omics contributions to personalized prediction of cardiovascular disease. Nat. Commun. 17, (2026).

19. Xie, R. et al. Improved sex-specific cardiovascular risk prediction with multi-omics data in people with type 2 diabetes. Cardiovasc. Diabetol. 25, 25 (2025).

20. Sritharan, H. P., Nguyen, H., Allahwala, U. K. & Bhindi, R. Machine Learning for Cardiovascular Risk Prediction: A Practical Primer for Clinicians. Heart Lung Circ. 35, 598–605 (2026).

21. Singh, H., Mhasawade, V. & Chunara, R. Generalizability challenges of mortality risk prediction models: A retrospective analysis on a multi-center database. PLOS digital health 1, e0000023 (2022).

22. Bild, D. E. et al. Multi-Ethnic Study of Atherosclerosis: Objectives and design. Am. J. Epidemiol. 156, 871–881 (2002).

23. Chen, Z.-Z. et al. Nontargeted and Targeted Metabolomic Profiling Reveals Novel Metabolite Biomarkers of Incident Diabetes in African Americans. Diabetes 71, 2426–2437 (2022).

24. Dean, D. A. et al. A systematic assessment of the association of polysomnographic indices with blood pressure: the Multi-Ethnic Study of Atherosclerosis (MESA). Sleep 38, 587–96 (2015).

25. Wyatt, S. B. et al. A community-driven model of research participation: the Jackson Heart Study Participant Recruitment and Retention Study. Ethn. Dis. 13, 438–55 (2003).

26. Wilson, J. G. et al. Study design for genetic analysis in the Jackson Heart Study. Ethn. Dis. 15, S6–30–37 (2005).

27. Fuhrman, B. J. et al. Assay Reproducibility and Interindividual Variation for 15 Serum Estrogens and Estrogen Metabolites Measured by Liquid Chromatography–Tandem Mass Spectrometry. Cancer Epidemiology, Biomarkers & Prevention 23, 2649–2657 (2014).

28. Paynter, N. P. et al. Metabolic Predictors of Incident Coronary Heart Disease in Women. Circulation 137, 841–853 (2018).

29. Zhang, Y. et al. Development and validation of a metabolite index for obstructive sleep apnea across race/ethnicities. Sci. Rep. 12, 21805 (2022).

30. Wilson, M. D., Ponzini, M. D., Taylor, S. L. & Kim, K. Imputation of Missing Values for Multi-Biospecimen Metabolomics Studies: Bias and Effects on Statistical Validity. Metabolites 12, 671 (2022).

31. Gogarten, S. M. et al. Genetic association testing using the GENESIS R/Bioconductor package. Bioinformatics 35, 5346–5348 (2019).

32. Purcell, S. et al. PLINK: A Tool Set for Whole-Genome Association and Population-Based Linkage Analyses. The American Journal of Human Genetics 81, 559–575 (2007).

33. Kurniansyah, N. et al. A multi-ethnic polygenic risk score is associated with hypertension prevalence and progression throughout adulthood. Nat. Commun. 13, 3549 (2022).

34. Kurniansyah, N. et al. Evaluating the use of blood pressure polygenic risk scores across race/ethnic background groups. Nat. Commun. 14, 3202 (2023).

35. Lambert, S. A. et al. The Polygenic Score Catalog as an open database for reproducibility and systematic evaluation. Nat. Genet. 53, 420–425 (2021).

36. Riley, R. D. et al. Penalization and shrinkage methods produced unreliable clinical prediction models especially when sample size was small. J. Clin. Epidemiol. 132, 88–96 (2021).

37. Tianqi Chen, Tong He & Michael Benesty. xgboost: Extreme Gradient Boosting. Preprint at (2026).

38. Kuhn, M. Building Predictive Models in R Using the caret Package. J. Stat. Softw. 28, (2008).

39. Mayer, M. shapviz: SHAP Visualizations. Preprint at https://github.com/modeloriented/shapviz (2025).

40. Hemani, G. et al. The MR-Base platform supports systematic causal inference across the human phenome. Elife 7, (2018).

41. Wald, A. The Fitting of Straight Lines if Both Variables are Subject to Error. The Annals of Mathematical Statistics 11, 284–300 (1940).

42. Sun, B. B. et al. Plasma proteomic associations with genetics and health in the UK Biobank. Nature 622, 329–338 (2023).

43. Huang, Y.-J. et al. A semi-empirical Bayes approach for calibrating weak instrumental bias in sex-specific Mendelian randomization studies. The American Journal of Human Genetics 112, 2213–2231 (2025).

44. Verma, A. et al. Diversity and scale: Genetic architecture of 2068 traits in the VA Million Veteran Program. Science (1979). 385, (2024).

45. Auton, A. et al. A global reference for human genetic variation. Nature 526, 68–74 (2015).

46. Szklarczyk, D. et al. The STRING database in 2023: protein–protein association networks and functional enrichment analyses for any sequenced genome of interest. Nucleic Acids Res. 51, D638–D646 (2023).

47. Wajid Jawaid. enrichR: Provides an R Interface to ‘Enrichr’. Preprint at https://github.com/wjawaid/enrichr (2025).

48. Kuleshov, M. V et al. Enrichr: a comprehensive gene set enrichment analysis web server 2016 update. Nucleic Acids Res. 44, W90–7 (2016).

49. Buniello, A. et al. Open Targets Platform: facilitating therapeutic hypotheses building in drug discovery. Nucleic Acids Res. 53, D1467–D1475 (2025).

50. Karczewski, K. J. & Snyder, M. P. Integrative omics for health and disease. Nat. Rev. Genet. 19, 299–310 (2018).

51. Hasin, Y., Seldin, M. & Lusis, A. Multi-omics approaches to disease. Genome Biol. 18, 83 (2017).

52. Rinschen, M. M., Ivanisevic, J., Giera, M. & Siuzdak, G. Identification of bioactive metabolites using activity metabolomics. Nat. Rev. Mol. Cell Biol. 20, 353–367 (2019).

53. Flint, A. C. et al. Effect of Systolic and Diastolic Blood Pressure on Cardiovascular Outcomes. N. Engl. J. Med. 381, 243–251 (2019).

54. Kim, M. et al. Proteomic Risk Score for Prediction of Incident Hypertension. Hypertension 83, (2026).

55. Drouard, G. et al. Multi-Omics Integration in a Twin Cohort and Predictive Modeling of Blood Pressure Values. OMICS 26, 130–141 (2022).

56. Vuori, M. et al. Value of Multiomics Over Clinical Risk Factors in Hypertension Prediction. Hypertension 83, (2026).

57. Keaton, J. M. et al. Genome-wide analysis in over 1 million individuals of European ancestry yields improved polygenic risk scores for blood pressure traits. Nat. Genet. 56, 778–791 (2024).

58. Mann, S. J. The Clinical Spectrum of Labile Hypertension: A Management Dilemma. The Journal of Clinical Hypertension 11, 491–497 (2009).

59. Kobori, H., Nangaku, M., Navar, L. G. & Nishiyama, A. The Intrarenal Renin-Angiotensin System: From Physiology to the Pathobiology of Hypertension and Kidney Disease. Pharmacol. Rev. 59, 251–287 (2007).

60. Potter, L. R., Yoder, A. R., Flora, D. R., Antos, L. K. & Dickey, D. M. Natriuretic peptides: their structures, receptors, physiologic functions and therapeutic applications. Handb. Exp. Pharmacol. 341–66 (2009) doi:10.1007/978-3-540-68964-5_15.

61. Salazar, J. et al. Neprilysin: A Potential Therapeutic Target of Arterial Hypertension? Curr. Cardiol. Rev. 16, 25–35 (2020).

62. Dhaun, N. et al. Role of Endothelin-1 in Clinical Hypertension. Hypertension 52, 452–459 (2008).

63. Lu, X. et al. The Role of CXC Chemokines in Cardiovascular Diseases. Front. Pharmacol. 12, (2022).

64. Rodríguez-Vita, J. et al. Angiotensin II Activates the Smad Pathway in Vascular Smooth Muscle Cells by a Transforming Growth Factor-β–Independent Mechanism. Circulation 111, 2509–2517 (2005).

65. Goumans, M.-J. & Ten Dijke, P. TGF-β Signaling in Control of Cardiovascular Function. Cold Spring Harb. Perspect. Biol. 10, (2018).

66. Tokolyi, A. et al. The contribution of genetic determinants of blood gene expression and splicing to molecular phenotypes and health outcomes. Nat. Genet. 57, 616–625 (2025).

67. Gioelli, N. et al. Neuropilin 1 and its inhibitory ligand mini-tryptophanyl-tRNA synthetase inversely regulate VE-cadherin turnover and vascular permeability. Nat. Commun. 13, 4188 (2022).

68. Kim, Y. T. et al. Highly secreted tryptophanyl tRNA synthetase 1 as a potential theranostic target for hypercytokinemic severe sepsis. EMBO Mol. Med. 16, 40–63 (2024).

69. Mohan, T., Deng, L. & Wang, B.-Z. CCL28 chemokine: An anchoring point bridging innate and adaptive immunity. Int. Immunopharmacol. 51, 165–170 (2017).

70. Yang, K. et al. CCL28 contributes to angiogenesis and cardiac repair through CCR10+ endothelial cells after myocardial infarction in male mice. Nat. Commun. 16, 9262 (2025).

71. Zhou, F. et al. Targeted multiplex proteomics for the development and validation of biomarkers in primary aldosteronism subtyping. Eur. J. Endocrinol. 191, 558–569 (2024).

72. Tsai, C. et al. Proteomic signatures to detect unilateral primary aldosteronism in hypertensive patients. Eur. J. Clin. Invest. 55, (2025).

73. Reel, P. S. et al. Machine learning for classification of hypertension subtypes using multi-omics: A multi-centre, retrospective, data-driven study. EBioMedicine 84, 104276 (2022).

