## Supplementary file 1 for "Multi-omics models for hypertension prediction using blood pressure PRS and omics scores"

**Table S2**[. Associations of blood pressure (BP) protein (ProtS) and metabolite (MetS) scores with corresponding BP traits in JHS	2](#_Toc238808858)

### **Table S1**. Polygenic risk score (PRS) development methods and source GWAS summary statistics by trait

| Trait | Development Method | Summary Statistics GWAS |
| --- | --- | --- |
| PRS_SBP | PRS-CSx (weighted sum of ancestry-specific SBP PRSs) | UKBB+ICBP, BBJ, COGENT |
| PRS_DBP | PRS-CSx (weighted sum of ancestry-specific DBP PRSs) | UKBB+ICBP, BBJ, COGENT |
| PRS_HTN | PRSice-2 (unweighted sum of multi-ethnic BP trait-specific PRSs) | UKBB (HTN), MVP (SBP, DBP) |

SBP: systolic blood pressure; DBP: diastolic blood pressure; HTN: hypertension; UKBB: UK Biobank; ICBP: International Consortium of Blood Pressure Genome-Wide Association Studies; MVP: Million Veteran Program; BBJ: BioBank Japan; COGENT: Cohorts for Genomics of Cardiometabolic Traits.

| Table S2. Associations of blood pressure (BP) protein (ProtS) and metabolite (MetS) scores with corresponding BP traits in JHS | | | |
| --- | --- | --- | --- |
|  | **Effect Size/Odds Ratio** | **P-value** | **Var Explained %** |
| Prots (N= 953) | | | |
| SBP | 0.72 | 1.96E-46 | 16.13 |
| DBP | 0.82 | 2.36E-43 | 17.81 |
| HTN | 2.05 | 2.50E-21 | 9.14 |
| MetS (N= 2469) | | | |
| SBP | 0.76 | 8.13E-18 | 2.42 |
| DBP | 0.46 | 5.77E-05 | 0.63 |
| HTN | 2.86 | 7.11E-25 | 3.82 |
| PRS (N= 3160) | | | |
| SBP | 2.18 | 4.27E-06 | 0.59 |
| DBP | 0.69 | 0.000135 | 0.44 |
| HTN | 1.26 | 7.59E-06 | 0.55 |

SBP: systolic blood pressure; DBP: diastolic blood pressure; HTN: hypertension

condition. Var: variance explained in association analysis in the test data.

N: sample size. Effect size was reported for SBP, DBP, with odds ratio for HTN.

| Table S3. Optimal hyperparameters selected for XGBoost prediction models | | | | | | |
| --- | --- | --- | --- | --- | --- | --- |
|  | **nrounds** | **max_depth** | **eta** | **min_child_weight** | **colsample** | **subsample** |
| Primary Analysis | | | | | | |
| Baseline | 100 | 3 | 0.01 | 8 | 0.8 | 0.8 |
| MetS | 300 | 3 | 0.01 | 8 | 0.8 | 0.8 |
| ProtS | 100 | 3 | 0.03 | 8 | 0.8 | 0.8 |
| PRS | 100 | 3 | 0.03 | 8 | 0.8 | 0.8 |
| All_scores | 100 | 3 | 0.05 | 5 | 0.8 | 0.8 |
| Sensitivity Analysis (+SBP, DBP) | | | | | | |
| Baseline | 200 | 3 | 0.01 | 8 | 0.8 | 0.8 |
| MetS | 100 | 3 | 0.01 | 10 | 0.8 | 0.8 |
| ProtS | 200 | 4 | 0.01 | 8 | 0.8 | 0.8 |
| PRS | 100 | 3 | 0.03 | 8 | 0.8 | 0.8 |
| All_scores | 300 | 4 | 0.01 | 5 | 0.8 | 0.8 |
| Sensitivity Analysis (ProtS and MetS developed using 80% MESA participants) | | | | | | |
| MetS | 300 | 3 | 0.01 | 10 | 0.8 | 0.8 |
| ProtS | 100 | 3 | 0.03 | 1 | 0.8 | 0.8 |

ProtS: protein risk scores; MetS: metabolite risk scores; PRS: polygenic risk scores.

| Table S4. Predictive performance for incident hypertension of Metabolite (MetS) and Protein (ProtS) Scores developed using 100% versus 80% of MESA participants | | | |
| --- | --- | --- | --- |
|  | **AUC** | **Calibration Intercept** | **Calibration Slope** |
| All MESA (n= N_MetS_ = 6345; N_ProtS_ = 5947) | | | |
| MetS | 0.69 (0.64 - 0.74) | 0.01 (-0.24 - 0.27) | 1.25 (0.92 – 1.66) |
| ProtS | 0.70 (0.66 - 0.75) | -0.05 (-0.31 - 0.21) | 1.05 (0.81 – 1.35) |
| 80% MESA (N_MetS_ = 5076; N_ProtS_ = 4758) | | | |
| MetS | 0.70 (0.65-0.74) | 0.07 (-0.18 – 0.34) | 1.34 (0.99 – 1.76) |
| ProtS | 0.70 (0.65-0.75) | -0.06 (-0.31 - 0.19) | 1.04 (0.81 – 1.34) |

AUC: area under the curve.

### **Table S5.** Causal proteins associated with blood pressure traits from Open Targets integration

| Gene | Significant outcome | MR effect (β) | FDR | Cardiovascular score | Leading drug (phase) | Tractability | Colocalized |
| --- | --- | --- | --- | --- | --- | --- | --- |
| **Established target: approved drugs with cardiovascular evidence** | | | | | | | |
| ADH1B | SBP_EUR | 0.237 | 2.44e-16 | 0.475 | FOMEPIZOLE (Approved) | SM, AB, PR | No |
| **AGT** | **HTN_EUR** | **0.132** | **3.74e-16** | **0.627** | **GSK-2586881 (PHASE 2)** | **SM, AB, PR, OC** | **Yes** |
| ASGR1 | SBP_EUR | -0.082 | 4.97e-06 | 0.078 | VUPANORSEN (PHASE 2) | SM, AB, PR, OC | No |
| SERPINE1 | SBP_EUR | 0.108 | 2.19e-05 | 0.314 | ALEPLASININ (PHASE 1) | SM, AB, PR | No |
| TGFB1 | DBP_EUR | 0.042 | 3.15e-04 | 0.418 | LUSPATERCEPT (Approved) | SM, AB, PR, OC | No |
| PCSK9 | DBP_EUR | -0.045 | 4.45e-04 | 0.724 | INCLISIRAN SODIUM (Approved) | SM, AB, PR, OC | No |
| PLAT | DBP_EUR | 0.102 | 1.66e-03 | 0.469 | MONTEPLASE (Approved) | SM, AB, PR, OC | No |
| ADH4 | DBP_EUR | -0.060 | 2.54e-03 | 0.257 | NITREFAZOLE (UNKNOWN) | SM, PR | No |
| F10 | DBP_EUR | -0.063 | 3.68e-03 | 0.559 | EDOXABAN (Approved) | SM, AB, PR, OC | No |
| CXCL8 | SBP_EUR | 0.077 | 4.65e-03 | 0.354 | ABX-IL8 (PHASE 2) | SM, AB, PR | No |
| TNFRSF9 | DBP_EUR | 0.047 | 5.07e-03 | 0.239 | UTOMILUMAB (PHASE 3) | SM, AB, PR | No |
| **Novel drug target: cardiovascular evidence but no approved drugs** | | | | | | | |
| NPPB | SBP_AFR, SBP_EUR*, HTN_EUR | -0.180 | 3.36e-34 | 0.385 | — | AB | No |
| APOA1 | SBP_EUR*, DBP_EUR | 0.308 | 1.61e-12 | 0.394 | — | SM, AB, PR | No |
| TNFRSF11B | DBP_EUR | -0.085 | 2.47e-12 | 0.114 | — | AB | No |
| SHBG | DBP_EUR | 0.085 | 6.27e-11 | 0.104 | — | SM, AB, PR | No |
| FES | DBP_EUR | -0.126 | 1.80e-10 | 0.576 | — | SM, AB, PR | No |
| WWP2 | SBP_EUR, DBP_EUR* | -0.098 | 3.45e-10 | 0.287 | — | SM, PR | No |
| LMOD1 | DBP_EUR | -0.081 | 2.17e-08 | 0.277 | — | SM | No |
| EDN1 | SBP_EUR | 0.055 | 9.20e-07 | 0.128 | — | AB, PR | No |
| MEGF9 | SBP_EUR | -0.038 | 9.42e-06 | 0.025 | — | AB, PR | No |
| ING1 | SBP_EUR | -0.234 | 2.19e-05 | 0.256 | — | PR | No |
| ZBTB17 | HTN_EUR | 0.537 | 3.34e-05 | 0.094 | — | SM, PR | No |
| C1QTNF1 | SBP_EUR | -0.048 | 3.61e-05 | 0.104 | — | AB | No |
| ICAM5 | HTN_EUR | -0.061 | 2.66e-04 | 0.155 | — | AB, PR | No |
| LAYN | HTN_EUR | -0.075 | 2.66e-04 | 0.147 | — | AB, PR | No |
| NADK | SBP_EUR | -0.039 | 3.50e-04 | 0.291 | — | SM, PR | No |
| **FGF5** | **SBP_AFR, SBP_EUR*** | **0.052** | **4.49e-04** | **0.548** | **—** | **AB** | **Yes** |
| MAN2B2 | DBP_EUR | -0.020 | 5.61e-04 | 0.005 | — | AB, PR | No |
| ADAMTS4 | DBP_EUR | 0.073 | 9.08e-04 | 0.061 | — | SM, AB, PR | No |
| WFDC12 | SBP_EUR | -0.021 | 1.32e-03 | 0.001 | — | AB | No |
| UMOD | DBP_EUR | 0.014 | 1.64e-03 | 0.553 | — | AB | No |
| CDH5 | HTN_EUR | -0.096 | 1.81e-03 | 0.329 | — | AB | No |
| SORBS1 | HTN_EUR | -0.369 | 2.06e-03 | 0.092 | — | AB, PR | No |
| KRT18 | SBP_EUR | -0.177 | 2.08e-03 | 0.080 | — | PR | No |
| HRC | HTN_EUR | 0.155 | 2.68e-03 | 0.062 | — | AB | No |
| NTproBNP | SBP_AFR*, HTN_AFR | -0.077 | 4.38e-03 | 0.385 | — | AB | No |
| ERP44 | DBP_EUR | 0.153 | 4.59e-03 | 0.101 | — | SM, AB, PR | No |
| IL16 | DBP_EUR | 0.013 | 5.06e-03 | 0.097 | — | AB, PR | No |
| VSNL1 | DBP_EUR | -0.031 | 5.10e-03 | 0.087 | — | PR | No |
| IGFBP7 | DBP_EUR | -0.037 | 5.26e-03 | 0.607 | — | AB, PR | No |
| **Repurposing candidate: approved drugs for non-cardiovascular indications** | | | | | | | |
| ICAM3 | SBP_EUR*, HTN_EUR | -0.031 | 3.50e-04 | 0.000 | ICM3 (UNKNOWN) | AB, PR | No |
| FLT3 | DBP_EUR | -0.103 | 9.12e-04 | 0.000 | QUIZARTINIB DIHYDROCHLORIDE (Approved) | SM, AB, PR, OC | No |
| COL2A1 | SBP_EUR | 0.025 | 3.87e-03 | 0.000 | OCRIPLASMIN (Approved) | SM, AB, PR, OC | No |
| **Novel finding: no prior cardiovascular evidence and no approved drugs** | | | | | | | |
| TCTN3 | SBP_EUR | 0.028 | 3.23e-11 | 0.000 | — | AB, PR | No |
| MXRA8 | SBP_EUR*, DBP_EUR | 0.223 | 9.42e-06 | 0.000 | — | AB, PR | No |
| VSIG2 | DBP_EUR | 0.061 | 2.02e-05 | 0.000 | — | AB | No |
| PLA2G1B | SBP_EUR*, DBP_EUR | -0.222 | 2.19e-05 | 0.000 | — | SM, AB, PR | No |
| **WARS** | **SBP_EUR*, DBP_EUR** | **0.167** | **2.19e-05** | **0.000** | **—** | **—** | **Yes** |
| **CCL28** | **DBP_EUR** | **0.198** | **3.17e-05** | **0.000** | **—** | **AB** | **Yes** |
| ANGPTL2 | HTN_EUR | 0.486 | 3.34e-05 | 0.000 | — | AB | No |
| NCAN | SBP_EUR*, DBP_EUR | -0.071 | 9.99e-05 | 0.000 | — | AB, PR | No |
| REG1B | HTN_EUR | 0.073 | 2.60e-04 | 0.000 | — | AB | No |
| AGER | DBP_EUR | 0.065 | 3.15e-04 | 0.000 | — | — | No |
| ANXA2 | DBP_EUR | -0.024 | 3.15e-04 | 0.000 | — | SM, AB, PR | No |
| PADI2 | DBP_EUR | 0.079 | 5.61e-04 | 0.000 | — | SM, AB, PR | No |
| PTPRF | SBP_EUR | 0.115 | 8.21e-04 | 0.000 | — | SM, AB, PR | No |
| ACAN | HTN_EUR | 0.146 | 1.72e-03 | 0.000 | — | AB | No |
| LTBP2 | HTN_EUR | -0.154 | 1.72e-03 | 0.000 | — | AB | No |
| SLC16A1 | HTN_EUR | -0.308 | 1.72e-03 | 0.000 | — | — | No |
| GFER | HTN_EUR | -0.264 | 1.72e-03 | 0.000 | — | — | No |
| DLL1 | HTN_EUR | -0.141 | 1.81e-03 | 0.000 | — | — | No |
| STC1 | SBP_EUR | -0.125 | 2.08e-03 | 0.000 | — | AB, PR | No |
| CES2 | HTN_EUR | -0.243 | 2.38e-03 | 0.000 | — | SM, AB, PR | No |
| CCDC50 | HTN_EUR | -0.236 | 2.43e-03 | 0.000 | — | PR | No |
| PTPRR | DBP_EUR | -0.209 | 2.59e-03 | 0.000 | — | AB, PR | No |
| GUSB | HTN_EUR | 0.136 | 2.63e-03 | 0.000 | — | SM, AB, PR | No |
| DDAH1 | SBP_EUR | 0.046 | 2.79e-03 | 0.000 | — | SM, AB, PR | No |
| PBXIP1 | DBP_EUR | 0.173 | 2.92e-03 | 0.000 | — | PR | No |
| HMOX2 | SBP_AFR*****, SBP_EUR | 0.147 | 3.76e-03 | 0.000 | — | SM, AB, PR | No |
| ERN1 | SBP_AFR | 0.163 | 3.76e-03 | 0.000 | — | SM, AB, PR | No |
| NELL1 | SBP_EUR | 0.031 | 3.86e-03 | 0.000 | — | AB, PR | No |
| CEBPB | DBP_EUR | 0.148 | 3.87e-03 | 0.000 | — | PR | No |
| AKT3 | DBP_EUR | -0.136 | 4.07e-03 | 0.000 | — | — | No |
| CCER2 | SBP_EUR | -0.070 | 4.65e-03 | 0.000 | — | — | No |
| SLC9A3R2 | SBP_EUR | 0.066 | 4.65e-03 | 0.000 | — | — | No |
| ENTPD5 | DBP_AFR | -0.079 | 4.70e-03 | 0.000 | — | AB, PR | No |
| PVR | DBP_EUR | -0.014 | 5.26e-03 | 0.000 | — | SM, AB, PR | No |

Bold rows indicate proteins with colocalization evidence (PP.H4 > 0.80).

MR effect size of primary outcome (indicated by *) was selected based on smallest FDR.

Outcomes: systolic (SBP) and diastolic pressure (DBP), hypertension (HTN) and essential HTN (ESS.HTN).

Ancestry groups: AFR: African; EUR: European.
OT: Open Targets cardiovascular association score (range 0-1).
Tractability: SM: small molecule; AB: antibody; PR: PROTAC; OC: other clinical.

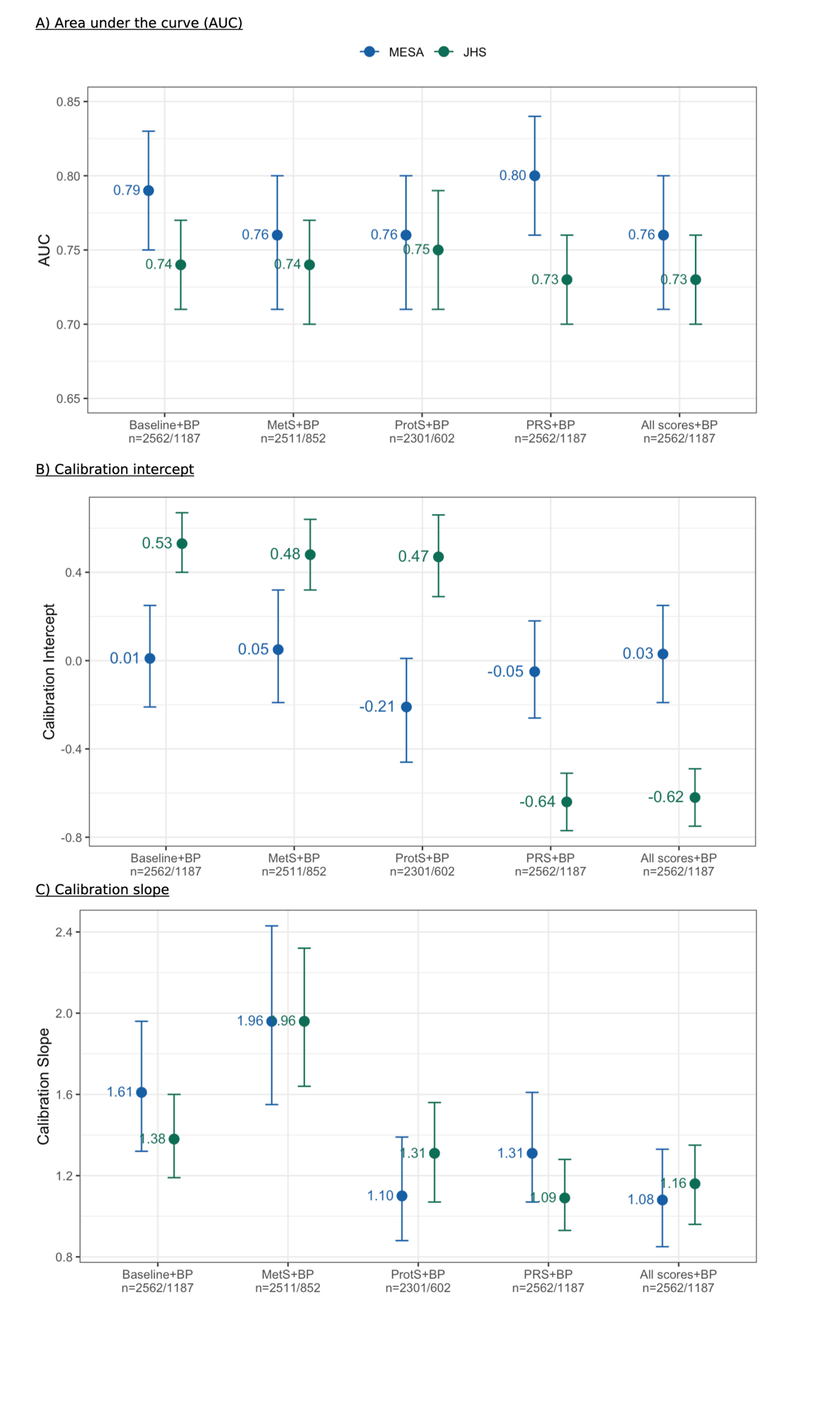

**Figure S1.** Discrimination and calibration of multi-omics XGBoost models with blood pressure (BP) measurements (systolic - SBP and diastolic - DBP) for incident hypertension prediction across MESA internal and JHS external validation cohorts in sensitivity analysis. A) Area under the curve (AUC), B) calibration intercept, and C) calibration slope for five progressively richer models with BP traits (SBP and DBP) as additional features: baseline clinical covariates, metabolomic scores (MetS), proteomic scores (ProtS), polygenic risk scores (PRS), and all omics scores combined. Models were validated on the held-out 20% MESA test set (blue), and externally validated using the full JHS cohort (green). Points represent point estimates and error bars represent 95% bootstrap confidence intervals. Sample sizes for each model are reported as n= n(MESA)/n(JHS).

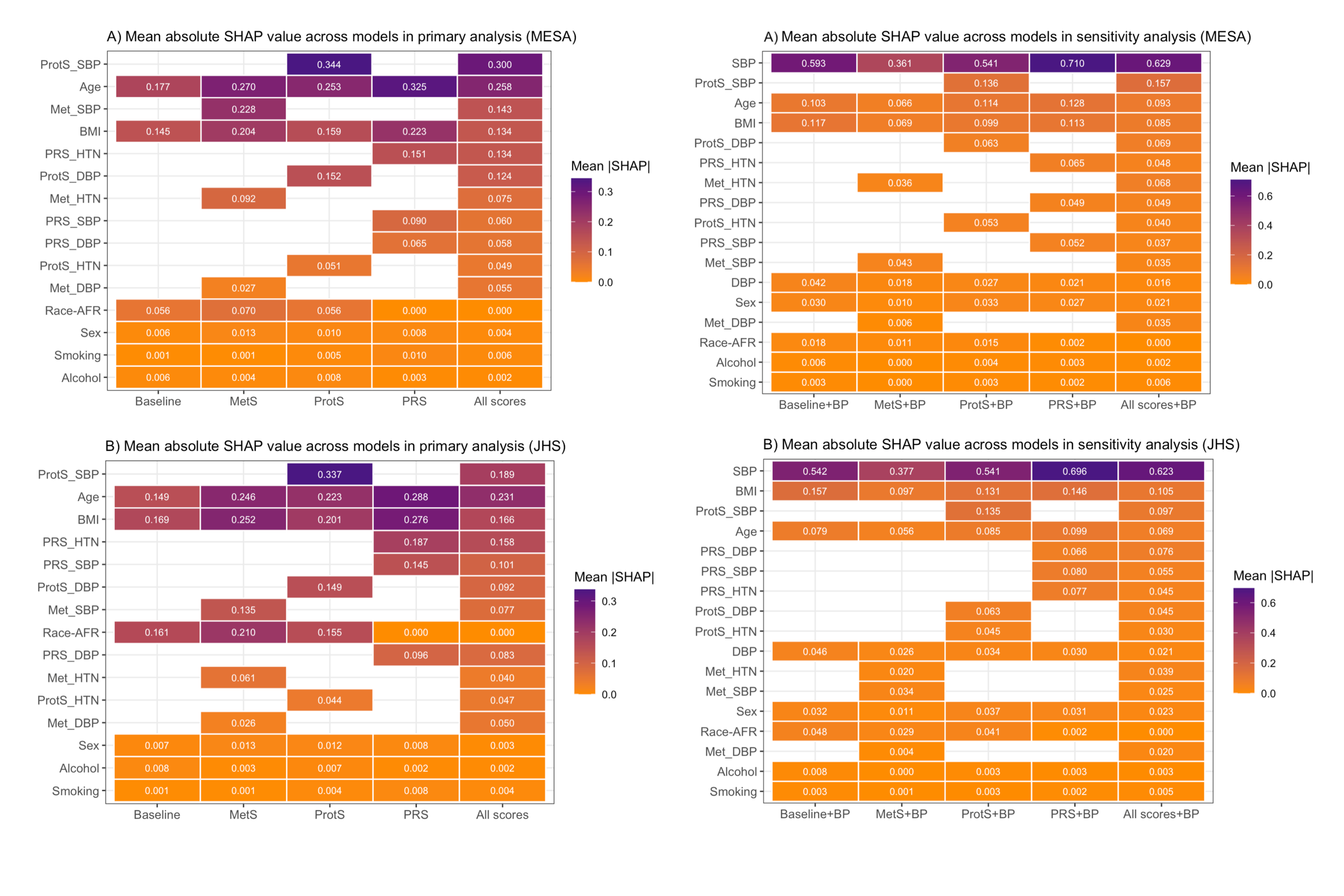

**Figure S2.** Feature importance across XGBoost models in primary analysis for MESA and JHS. Mean absolute SHAP (SHapley Additive exPlanations) values across five models in A) MESA internal validation, B) JHS external validation. Features are ordered by overall importance. Empty cells indicate features not included in the respective model.

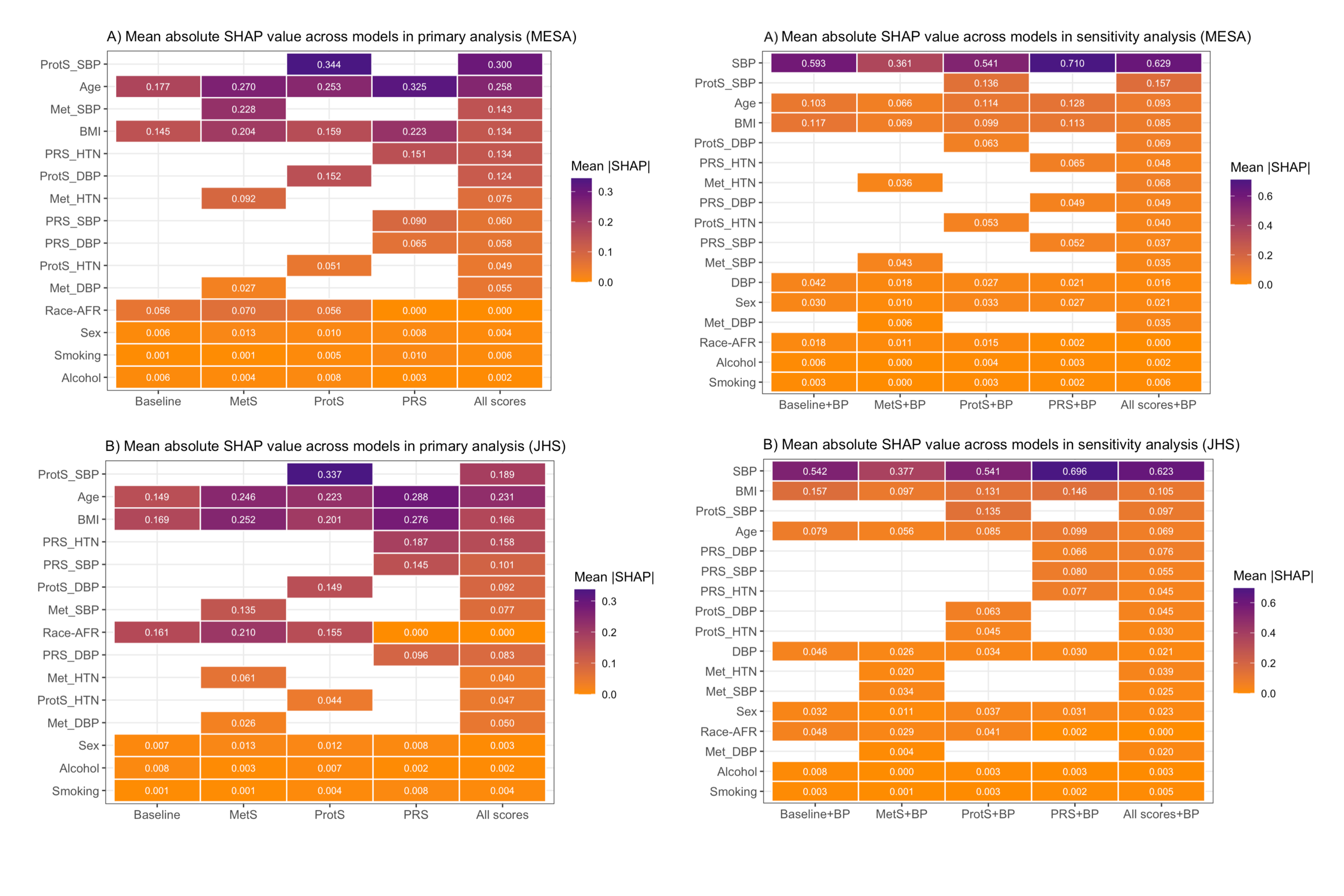

**Figure S3.** Feature importance across XGBoost models in sensitivity analysis including measured systolic (SBP) and diastolic blood pressure (DBP) for MESA and JHS. Mean absolute SHAP (SHapley Additive exPlanations) values across five models in A) MESA internal validation, B) JHS external validation, Features are ordered by overall importance. Empty cells indicate features not included in the respective model.

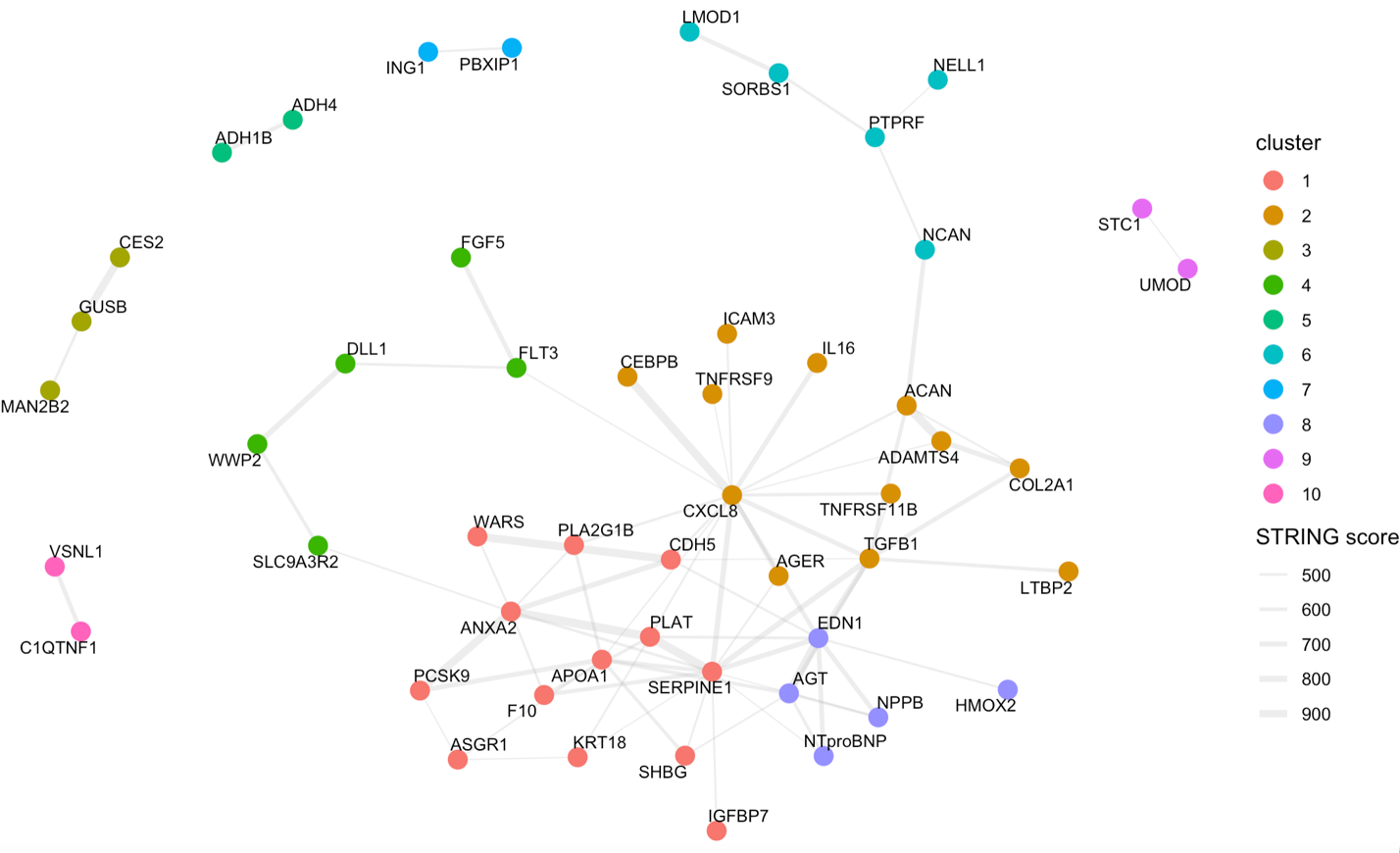

**Figure S4.** Protein-protein interaction network of Mendelian randomization (MR) significant proteins associated with blood pressure traits. Node colors represent functional clusters identified using the STRING database. Edge thickness reflects STRING interaction confidence score.

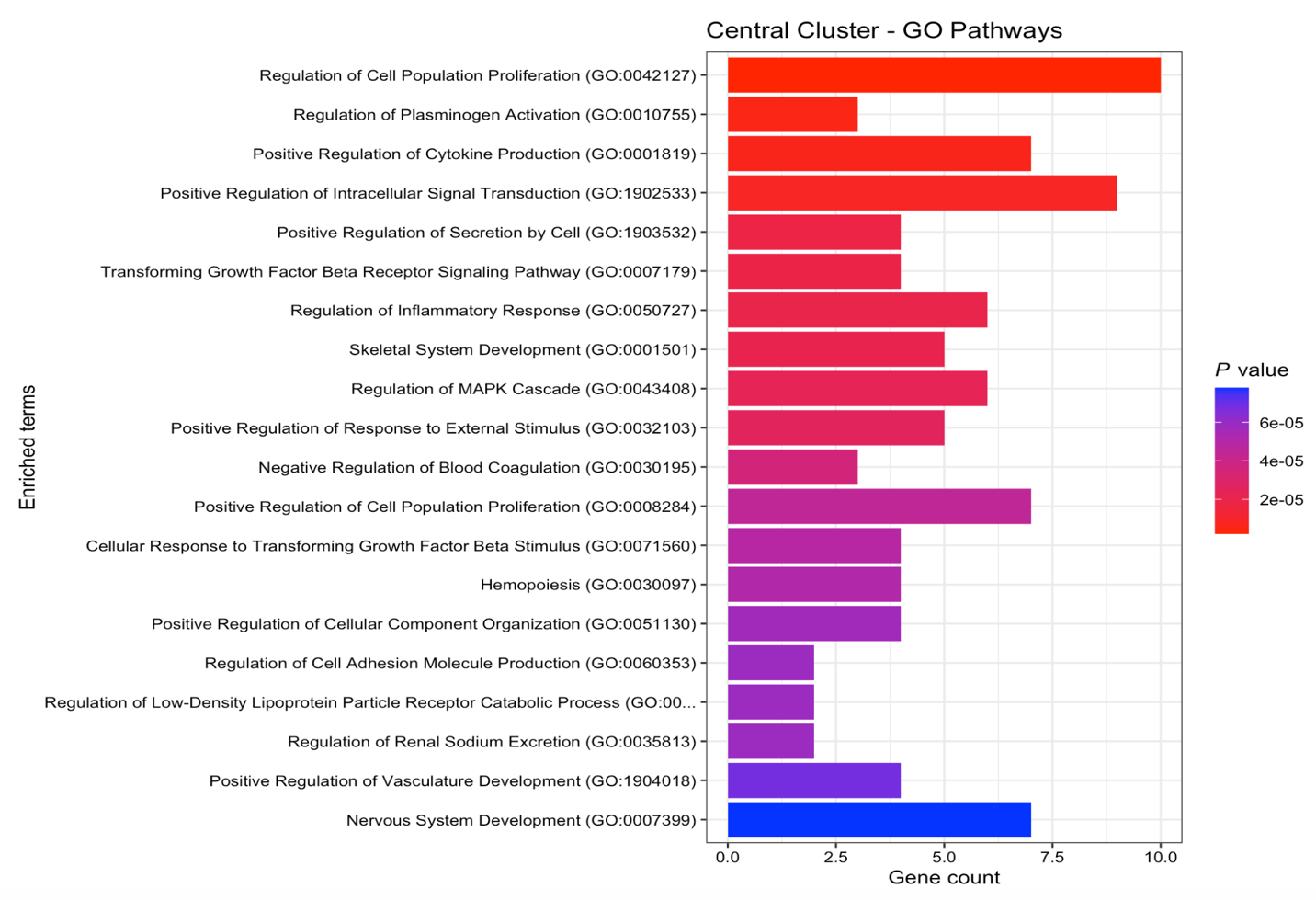

**Figure S5.** Top 20 Gene Ontology biological process enrichment of the central STRING interaction cluster of Mendelian randomization significant blood pressure proteins. Bars represent gene count per enriched term, colored by nominal p-value from Enrichr analysis (red = most significant, blue = least significant).
